# Omicron BA.1/BA.2 infections in triple-vaccinated individuals enhance a diverse repertoire of mucosal and blood immune responses

**DOI:** 10.1101/2023.01.28.23285084

**Authors:** Hailey Hornsby, Alexander R. Nicols, Stephanie Longet, Chang Liu, Adriana Tomic, Adrienn Angyal, Barbara Kronsteiner, Jessica K. Tyerman, Tom Tipton, Peijun Zhang, Marta Gallis Ramalho, Piyada Supasa, Muneeswaran Selvaraj, Priyanka Abraham, Isabel Neale, Mohammad Ali, Natalie A. Barratt, Jeremy M. Nell, Lotta Gustafsson, Scarlett Strickland, Irina Grouneva, Timothy Rostron, Shona C. Moore, Luisa M. Hering, Susan L. Dobson, Sagida Bibi, Juthathip Mongkolsapaya, Teresa Lambe, Dan Wootton, Victoria Hall, Susan Hopkins, Tao Dong, Eleanor Barnes, Gavin Screaton, Alex Richter, Lance Turtle, Sarah L. Rowland-Jones, Miles Carroll, Christopher J.A. Duncan, Paul Klenerman, Susanna J. Dunachie, Rebecca P. Payne, Thushan I. de Silva, the PITCH consortium

**Author notes:** Corresponding authors: Thushan de Silva, Department of Infection, Immunity and Cardiovascular Disease, Medical School, University of Sheffield, S10 2RX, Sheffield, UK;, Paul Klenerman, Peter Medawar Building for Pathogen Research, University of Oxford, Oxford OX1 3SY, UK. These authors contributed equally (Hailey Hornsby, Alexander R. Nicols, Stephanie Longet, Chang Liu). Senior authors (Alex Richter, Lance Turtle, Sarah L. Rowland-Jones, Miles Carroll, Christopher J.A. Duncan, Paul Klenerman, Susanna J. Dunachie, Rebecca P. Payne, Thushan I. de Silva). Full list of consortium authors can be found in supplementary information.

## Abstract

Pronounced immune escape by the SARS-CoV-2 Omicron variant has resulted in large numbers of individuals with hybrid immunity, generated through a combination of vaccination and infection. Based primarily on circulating neutralizing antibody (NAb) data, concerns have been raised that omicron breakthrough infections in triple-vaccinated individuals result in poor induction of omicron-specific immunity, and that a history of prior SARS-CoV-2 in particular is associated with profound immune dampening. Taking a broader and comprehensive approach, we characterized mucosal and blood immunity to both spike and non-spike antigens following BA.1/BA.2 infections in triple mRNA-vaccinated individuals, with and without a history of previous SARS-CoV-2 infection. We find that the majority of individuals increase BA.1/BA.2/BA.5-specific NAb following infection, but confirm that the magnitude of increase and post-omicron titres are indeed higher in those who were infection-naive. In contrast, significant increases in nasal antibody responses are seen regardless of prior infection history, including neutralizing activity against BA.5 spike. Spike-specific T cells increase only in infection-naive vaccinees; however, post-omicron T cell responses are still significantly higher in previously-infected individuals, who appear to have maximally induced responses with a CD8+ phenotype of high cytotoxic potential after their 3^rd^ mRNA vaccine dose. Antibody and T cell responses to non-spike antigens also increase significantly regardless of prior infection status, with a boost seen in previously-infected individuals to immunity primed by their first infection. These findings suggest that hybrid immunity induced by omicron breakthrough infections is highly dynamic, complex, and compartmentalised, with significant immune enhancement that can help protect against COVID-19 caused by future omicron variants.

## Introduction

Since its initial description in November 2021, the B.1.1.529 (omicron) variant of severe acute respiratory syndrome coronavirus 2 (SARS-CoV-2) has rapidly spread throughout the world (1,2). Several omicron-lineage viruses have since emerged, with waves dominated initially by BA.1 and BA.2 variants, followed by BA.4/5, and more recently by combinations of omicron variants such as BA.2.75, BQ.1 and XBB (3). The unprecedented number of spike mutations in omicron viruses has led to considerable immune escape from vaccine- and infection-induced immunity (4,5). Vaccine effectiveness against symptomatic SARS-CoV-2 infection with B.1.1.529 after a third BNT162b2 mRNA vaccine dose is estimated to be 67.2% at 2 - 4 weeks, falling to 45.7% after 10 weeks (6). As a result, a large number of individuals in highly vaccinated populations now have so-called hybrid immunity, generated through a combination of vaccination and infection. A multi-region cohort study in Switzerland estimated that at least 51% of the population had hybrid immunity by July 2022, with 85% having some degree of omicron-specific antibody immunity (7).

Until widespread circulation of omicron, individuals with hybrid immunity were primarily those who were infected during SARS-CoV-2 B.1.1.7/alpha or pre-alpha (‘ancestral’) waves, prior to commencing their vaccine courses. These ‘previously-infected’ individuals have higher spike-specific serum antibody and T cell responses after each vaccine dose compared to infection-naive vaccinees (8–10). Hybrid immunity generated by post-vaccination infections may be quantitatively and qualitatively different from responses seen in individuals who experienced SARS-CoV-2 infection before receiving a vaccination course. This may be due to differences in the priming SARS-CoV-2 exposure or lower antigenic exposure during the attenuated disease course of omicron viruses; although it is difficult to tease apart the contributions of viral phenotype change from those of pre-existing immunity (11). Recent reports have suggested that omicron-specific immunity generated by breakthrough infections may be muted in triple-vaccinated individuals, with those with a history of prior SARS-CoV-2 having a particularly poor response due to immune ‘imprinting’ from their previous infection (12,13).

We undertook comprehensive profiling of circulating and mucosal immunity before and after omicron BA.1 or BA.2 infections in a multi-site cohort of triple-vaccinated healthcare workers in the United Kingdom, stratified by those with a history of prior SARS-CoV-2 infection and those who were infection-naive. We find that whilst SARS-CoV-2-specific neutralizing antibody (NAb) responses to omicron infection are indeed lower in those with prior SARS-CoV-2, consistent with recent observations, omicron-specific neutralizing activity nevertheless increases significantly in most individuals. Spike-specific T cell responses also increase only in those with no prior history of SARS-CoV-2, although the magnitude of these responses is still higher in previously-infected individuals after omicron infection. Furthermore, increases in secretory IgA responses in nasal mucosal lining fluid are seen post-omicron, regardless of prior SARS-CoV-2 history, as are antibody and T cell responses to non-spike targets. Our data demonstrate that previous SARS-CoV-2 infection history may modulate immune responses to spike upon omicron infection in vaccinated populations. However, omicron infection in triple-vaccinated individuals generally enhances immune responses to SARS-CoV-2 in both blood and mucosa and is likely to contribute to ongoing population immunity against COVID-19.

## Results

### Participants

94 individuals from four PITCH sites (Liverpool, Newcastle, Oxford and Sheffield) with SARS-CoV-2 infection occurring after three BNT162b2 mRNA vaccine doses were included in the study, of which 38 (40.4%) had a history of prior SARS-CoV-2 before commencing their vaccine course (Table 1). The median time from this first infection to the 3rd vaccine dose was 544 days (IQR 514-559), with all but one infection occurring prior to December 2020 when widespread circulation of B.1.1.7/alpha variants occurred (pre-alpha/ancestral). Previously-infected individuals were slightly older than naive healthcare workers (median age 48 vs 41, p=0.02, Table 1), with a higher proportion of female participants (84.2% vs. 58.9%, p=0.01). All individuals had received their 1st and 2nd vaccine doses a median of 9.6 weeks apart (IQR 8.9-10.9). Omicron infections occurred between 21st December 2021 and 17th May 2022, with sequence data available from 29% of individuals included in the study to confirm SARS-CoV-2 lineage (17 BA.1 and 11 BA.2). Assuming that infections from 20th March 2022 were likely to be caused by BA.2 (14), 62 (66%) infections were classified as probable or confirmed BA.1 (Table 1). Full details of included participants are provided in Table 1.

**Table 1.** Details of participants included in the study.

|  | SARS-CoV-2 naive | Previously-infected | *P value* |
| --- | --- | --- | --- |
| **Number** | 56 | 38 | - |
| **Median age in years (IQR)** | 41 (35-49.8) | 48 (39.8-55.0) | 0.02 |
| **Sex** |  |  |  |
| Female | 33 (58.9%) | 32 (84.2%) | 0.01 |
| Male | 23 (41.1%) | 6 (15.8%) |  |
| **Median time (days) from previous infection to 3rd mRNA vaccine dose (IQR)** | NA | 544 (514-559) | - |
| **Median time (days) from 3rd mRNA vaccine to omicron infection (IQR)** | 130 (88-167) | 146 (95-167) | 0.55 |
| **Median time (days) from 3rd mRNA vaccine dose to pre-omicron sample (IQR)** | 33 (28-49) | 30 (28-36) | 0.12 |
| **Median time (days) from omicron infection to post-omicron sample (IQR)** | 31 (28-34) | 30 (28-36) | 0.92 |
| **Omicron subvariant\*** |  |  |  |
| BA.1 (sequence confirmed) | 39 (8) | 23 (9) | - |
| BA.2 (sequence confirmed) | 17 (5) | 15 (6) | - |
| **Site** |  |  |  |
| Liverpool | 4 | 1 | - |
| Newcastle | 14 | 9 | - |
| Oxford | 22 | 7 | - |
| Sheffield | 16 | 21 | - |

\* Classified by sequence data or based on a cut-off date of 20th March 2022, when >90% of SARS-CoV-2 infections in the UK were due to BA.2; IQR = interquartile range;

### Peripheral blood and nasal immune responses prior to omicron infection

Following the 3rd mRNA vaccine dose, but prior to omicron infection, individuals with a history of previous SARS-CoV-2 infection (i.e. hybrid immunity) had significantly higher NAb titres against ancestral (p=0.006), BA.1 (p=0.04), BA.2 (p=0.005) and BA.5 (p=0.007) viruses than SARS-CoV-2 naive healthcare workers (Figure 1A). While plasma spike-specific IgG was equivalent in both groups, plasma spike-specific IgA was higher in previously-infected individuals to ancestral (p=0.002), BA.2 (p=0.01) and BA.5 (p=0.03) proteins (Figure 1B & 1C), as was plasma nucleocapsid-specific IgG (p<0.0001, Figure 1D). Only 13 of 38 (35.1%) previously-infected individuals had detectable plasma nucleocapsid-specific IgG, likely reflecting waning of anti-N IgG from initial SARS-CoV-2 infection, which occurred almost exclusively in pre-alpha waves. No significant differences between previously-infected and naive participants were seen in spike- or nucleocapsid-specific secretory IgA (sIgA) from nasal lining fluid (Figure 1E & 1F), with equivalent human angiotensin-converting enzyme 2 (ACE2) inhibiting activity in both groups against ancestral, BA.2 and BA.5 spike proteins (Figure 1G). Peripheral T cell responses were significantly higher in previously-infected individuals against spike S1, S2, and combined membrane and nucleocapsid peptide pools (Figures 1H-J), as previously demonstrated (9).

**Figure 1.**
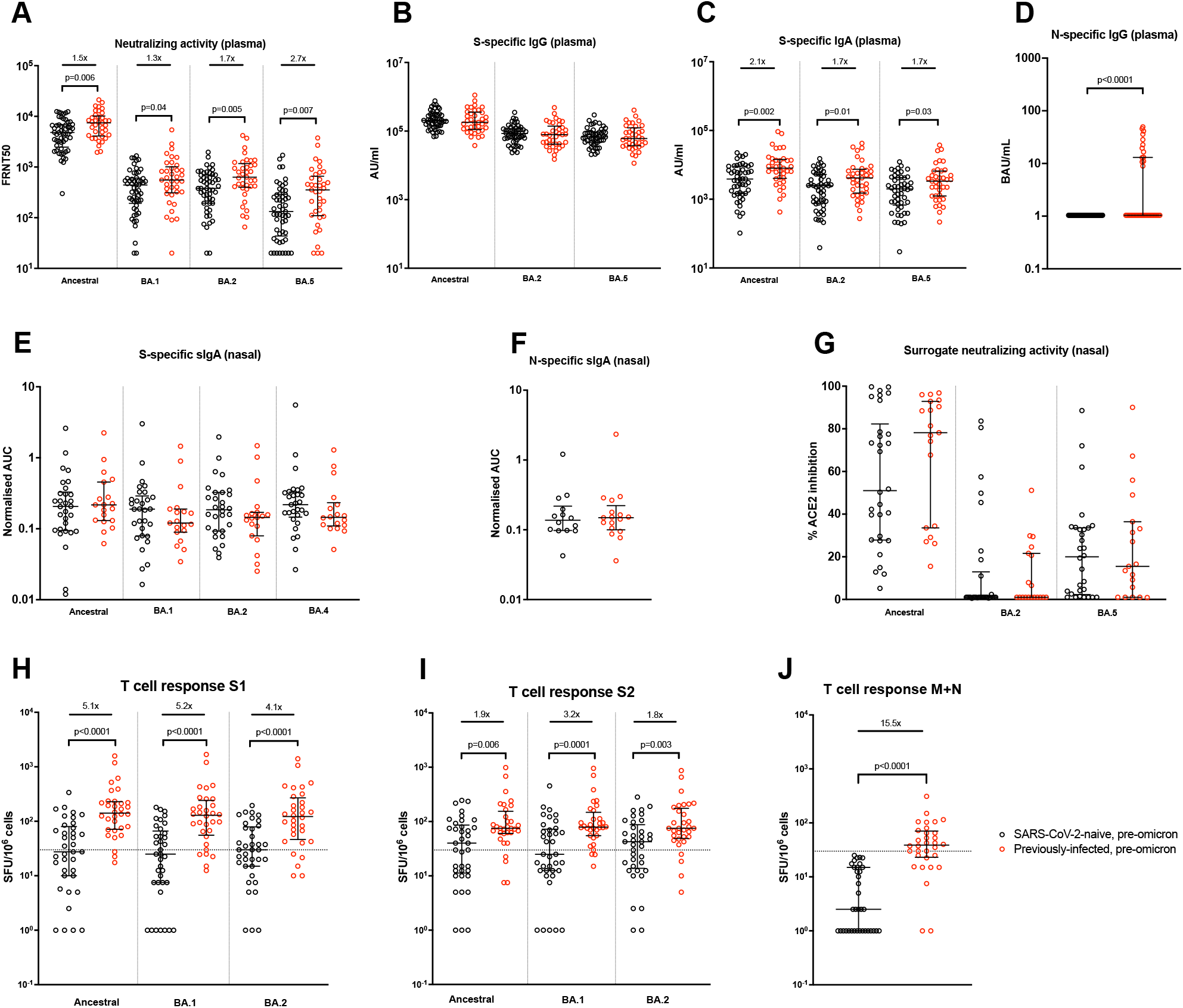
Comparison of immune responses prior to omicron infection in SARS-CoV-2-naive and previously-infected individuals. Samples were taken a median of 32 days (IQR 28-42.3) after 3rd mRNA dose. (A) Live-virus neutralizing activity of plasma against ancestral, BA.1, BA.2 and BA.5 viruses, expressed as the reciprocal of the dilution showing 50% reduction in focus forming units (FRNT50); SARS-CoV-2 spike-specific binding IgG (B) and IgA (C) in plasma against ancestral, BA.2 and BA.5 spike proteins (AU/mL = arbitrary antibody units/mL in MSD assay); (D) nucleocapsid-specific IgG in plasma, assessed by ELISA and expressed in WHO International units, BAU/mL; Secretory IgA (sIgA) in nasal lining fluid targeting (E) ancestral, BA.1, BA.2 and BA.4 spike proteins and (F) nucleocapsid-specific, expressed as area under the curve (AUC) normalised to total sIgA; (G) ability of nasal lining fluid to inhibit ACE2 binding to ancestral, BA.2 and BA.5 spike proteins, assessed by MSD assay; IFN-γ ELISpot responses to overlapping peptide pools representing the S1 (H) and S2 (I) spike subunits of ancestral, BA.1 and BA.2 viruses, and (J) a single pool containing peptides of both the ancestral membrane (M) and nucleocapsid (N) proteins. Results expressed as spot-forming units per million cells (SFU/106). The dashed line represents a positivity threshold of the mean + 2SD of the background response. Data shown with median and interquartile range. Median fold-difference between infection-naive and previously-infected individuals is displayed. All comparisons made with a Mann-Whitney U test. P values are displayed where <0.05.

### Impact of omicron infection on neutralizing and binding antibody responses

The median time from 3rd mRNA vaccine dose to omicron infection was 140 days (IQR 90-167, Table 1) and not significantly different between SARS-CoV-2 naive and previously-infected individuals (p=0.55). Following omicron infection, a significant increase in plasma ancestral virus neutralizing ability was seen in previously SARS-CoV-2 naive individuals (2.2-fold, p<0.0001), but not in previously-infected healthcare workers (1.0-fold, p=0.71, Figure 2A). A much greater increase in neutralizing activity was seen in the previously-naive group to omicron BA.1 (7.3-fold, p<0.0001), BA.2 (5.8-fold, p<0.0001) and BA.5 (8.1-fold, p<0.0001) viruses. Previously-infected individuals also had a boost in NAbs to omicron variants after infection, although to a lesser extent (BA.1, 1.7-fold, p=0.002; BA.2, 1.4-fold, p=0.002; BA.5, 1.6-fold, p=0.007). The post-infection neutralizing titres to all variants tested were significantly higher in the previously-naive individuals than in those with a history of prior SARS-CoV-2 (Figure 2A). Significant heterogeneity was evident in individual plasma NAb trajectories to BA.1 and BA.2 (Figure 2B & 2C), with antibody responders and non-responders seen in both groups. A NAb increase (defined as a fold difference >1.0) was seen in 49/53 naive individuals to BA.1 and 50/53 to BA.2, compared to BA.1 and BA.2 NAb increases in 26/37 previously-infected individuals.

**Figure 2.**
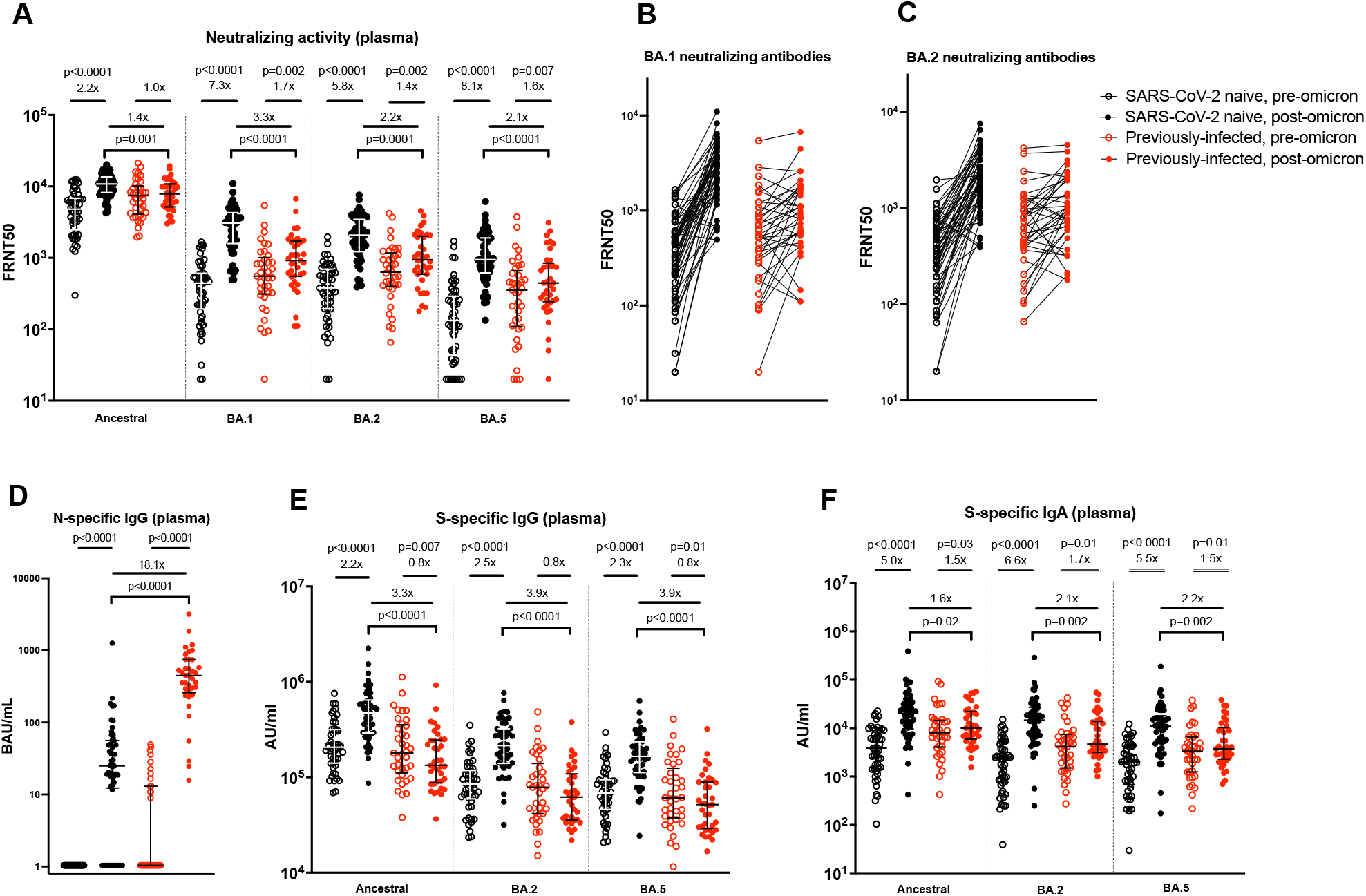
Impact of omicron infection on plasma neutralizing and binding antibodies in SARS-CoV-2-naive and previously-infected individuals. (A) Live-virus neutralizing activity of plasma against ancestral, BA.1, BA.2 and BA.5 viruses, expressed as the reciprocal of the dilution showing 50% reduction in focus forming units (FRNT50); Pair-wise depiction of pre- and post-omicron FRNT as individual participant trajectories for BA.1 (B) and BA.2 (C) neutralizing antibodies; (D) Nucleocapsid-specific IgG in plasma, assessed by ELISA and expressed in WHO International units, BAU/mL; SARS-CoV-2 spike-specific binding IgG (E) and IgA (F) in plasma against ancestral, BA.2 and BA.5 spike proteins (AU/mL = arbitrary antibody units/mL in MSD assay); Data shown with median and interquartile range. Median fold-change from pre-to post-infection samples is displayed. Statistical comparisons of paired pre- and post-infection samples done with Wilcoxon signed-rank test, and between post-infection levels in previously-infected and SARS-CoV-2 naive individuals using the Mann-Whitney U test. P values are displayed where <0.05. Responses were evaluated in 53 SARS-CoV-2-naive and 37 previously-infected individuals for whom samples were available. Post-omicron samples were taken a median of 30 days after infection.

Plasma anti-nucleocapsid IgG increased in both groups following infection, although in contrast to the neutralization data, post-infection levels were significantly higher in the previously-infected individuals than in those who were SARS-CoV-2 naive prior to omicron infection (p<0.0001, Figure 2D). All previously-infected individuals now had detectable anti-nucleocapsid IgG, compared to 41/53 (77.4%) of previously-naive healthcare workers.

Anti-spike binding IgG levels in plasma followed a similar pattern to the NAb data in previously-naive individuals, with an increase following infection seen to ancestral, BA.2 and BA.5 spike proteins, and post-infection levels now significantly higher in these individuals compared to those with prior SARS-CoV-2 (Figure 2E). In the previously-infected group, anti-spike IgG levels to all proteins were slightly lower following infection compared to their post-dose 3 level (ancestral, 0.8-fold, p=0.007; BA.2, 0.8-fold, p=0.09; BA.5, 0.8-fold, p=0.01), although this comparison does not account for any waning of responses that occurred after dose 3 but prior to omicron infection. In contrast, plasma anti-spike binding IgA increased following infection in both groups (Figure 2F), although again to a greater extent in previously-naive (ancestral, 5.0-fold, p<0.0001; BA.2, 6.6-fold, p<0.0001; BA.5, 5.5-fold, p<0.0001) compared to previously-infected individuals (ancestral, 1.5-fold, p=0.03; BA.2, 1.7-fold, p=0.01; BA.5, 1.5-fold, p=0.01). Similar findings were observed when stratifying participants by likely BA.1 or BA.2 infections (Figure S1).

### Nasal secretory IgA and functional antibody responses following omicron infection

Nasal spike-specific sIgA increased significantly following omicron infection in both SARS-CoV-2 naive and previously-infected individuals (Figure 3A). The extent of increase was similar across the different proteins tested; 11.4-fold to 16.5-fold increases in SARS-CoV-2 naive individuals and 10.6-fold to 14.8-fold increases in previously-infected individuals, with no statistically significant differences in post-infection levels between the two groups. Similarly, nasal nucleocapsid-specific sIgA also increased significantly following omicron infection in both SARS-CoV-2 naive (3.1-fold, p=0.003) and previously-infected (2.9-fold, p=0.0002) healthcare workers (Figure 3B). The ability of nasal lining fluid to inhibit ACE2 binding to ancestral spike protein (surrogate neutralization assay) was significantly increased post-infection in previously-naive individuals (p=0.003), but not in those with a prior history of SARS-CoV-2 (p=0.95; Figure 3C). In contrast, significant increases in inhibition of ACE2 binding to both BA.2 and BA.5 spike proteins was seen in both groups (Figure 3C).

**Figure 3.**
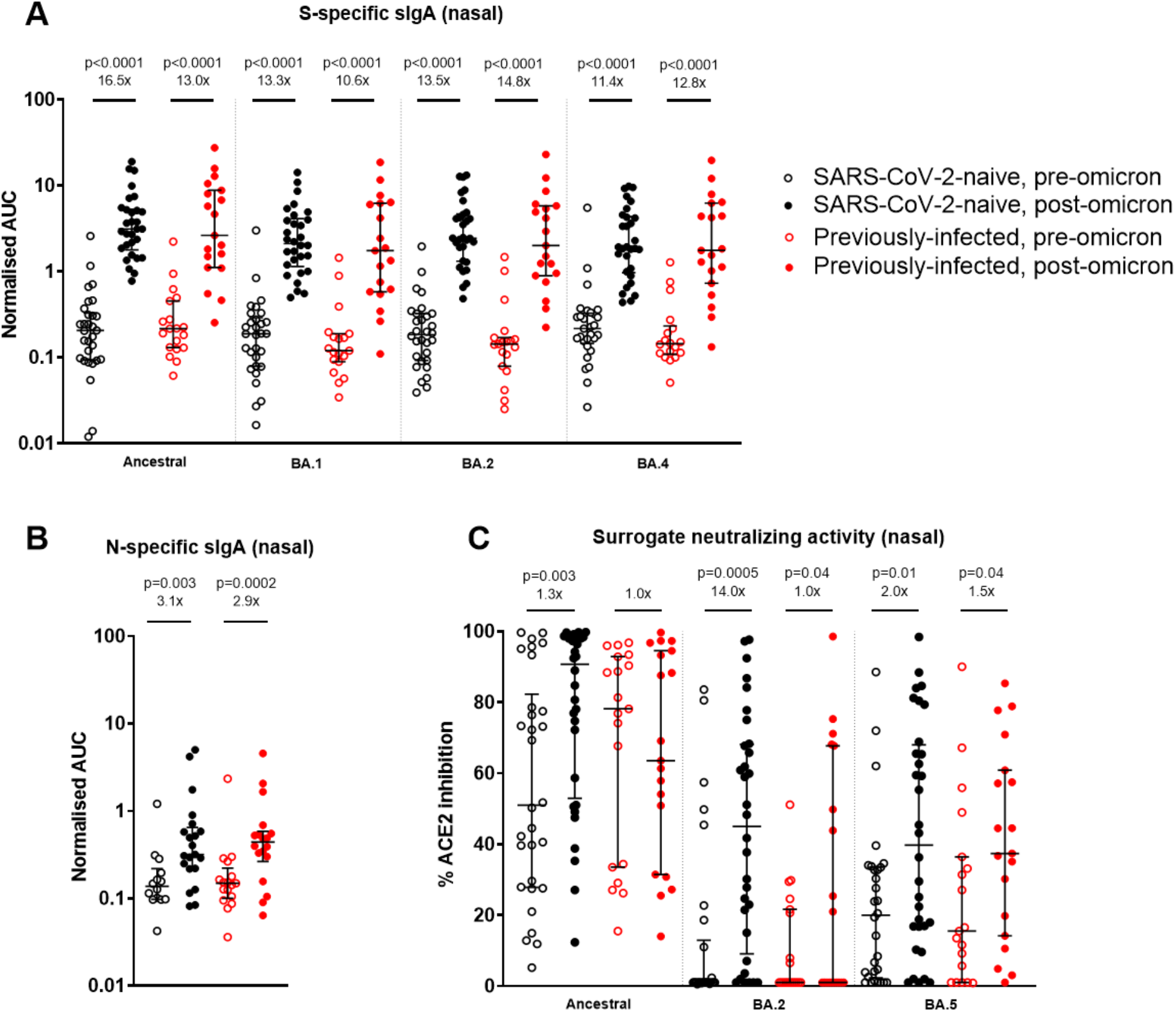
Impact of omicron infection on secretory IgA and ability to inhibit ACE2 binding to spike proteins in nasal lining fluid in SARS-CoV-2-naive and previously-infected individuals. Secretory IgA (sIgA) in nasal lining fluid targeting (A) ancestral, BA.1, BA.2 and BA.4 spike proteins and (B) nucleocapsid protein, expressed as area under the curve (AUC) normalised to total sIgA; (C) ability of nasal lining fluid to inhibit ACE2 binding to ancestral, BA.2 and BA.5 spike proteins, assessed by MSD assay. Data shown with median and interquartile range. Median fold-change from pre-to post-infection samples is displayed. Statistical comparisons of paired pre- and post-infection samples done with Wilcoxon signed-rank test, and between post-infection levels in previously-infected and SARS-CoV-2 naive individuals using the Mann-Whitney U test. P values are displayed where <0.05. Responses were evaluated in 32 SARS-CoV-2-naive and 19 previously-infected individuals for whom samples were available.

### Impact of omicron infection on spike and non-spike T cell responses

In SARS-CoV-2 naive individuals, a significant increase following omicron infection was seen in peripheral IFN-***γ*** ELISpot responses to peptide pools representing ancestral S1 (1.9-fold, p<0.0001) and S2 (1.8-fold, p=0.002), BA.1 S1 (2.7-fold, p<0.0001) and S2 (2.2-fold, p=0.0006), and BA.2 S1 (2.6-fold, p<0.0001) and S2 (1.8-fold, p=0.002; Figures 4A & 4B). No increase following omicron was seen in those with a previous SARS-CoV-2 infection, however in contrast to plasma neutralizing antibodies, post-omicron S1-specific T cell responses were still significantly higher than in previously-naive individuals for ancestral (p=0.003), BA.1 (p=0.004) and BA.2 (p=0.01) peptide pools (Figure 4A). In contrast to spike responses, T cell responses to a matrix and nucleocapsid peptide pool increased in both SARS-CoV-2 naive (11.8-fold, p<0.0001) and previously-infected (2.0-fold, p=0.0004) individuals, with post-omicron levels significantly higher in the latter group following a boost to previously primed T cells targeting these non-spike antigens (p=0.005, Figure 4C). Significant increases in ELISpot responses to peptide pools representing ancestral non-structural proteins (NSP)1-2 (2.4-fold, p=0.002), NSP4-6 (2.5-fold, p=0.02), NSP7-11 (1.6-fold, p=0.002) and NSP12 (1.6-fold, p=0.04) were seen in SARS-CoV-2 naive individuals after omicron infection (Figure S2), although overall responses in each pool remained low. Omicron infection induced significant increases to a pool of peptides corresponding to NSP3 amino acids 663-1945 in both naive (2.9-fold, p=0.0008) and previously-infected (2.6-fold, p=0.0006) individuals (Figure S2).

**Figure 4.**
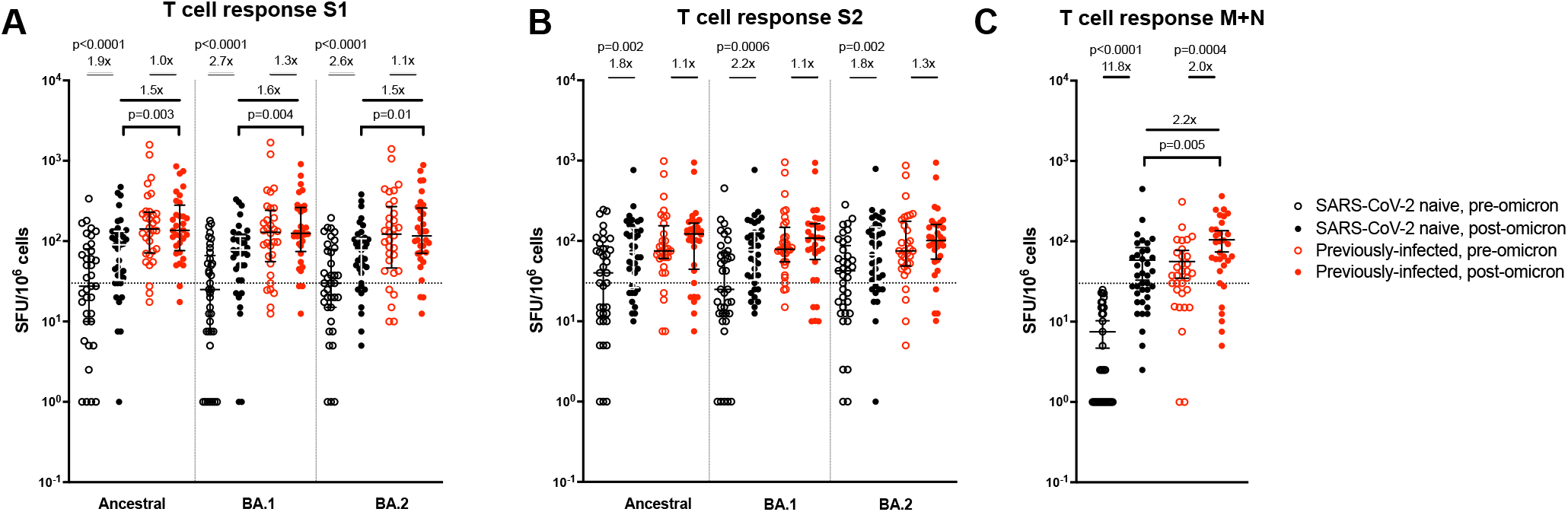
Impact of omicron infection on T cell response to SARS-CoV-2 proteins in SARS-CoV-2-naive and previously-infected individuals. IFN-**γ** ELISpot responses to overlapping peptide pools representing the S1 (A) and S2 (B) spike subunits of ancestral, BA.1 and BA.2 viruses, and (C) a single pool containing peptides of both the ancestral membrane (M) and nucleocapsid (N) proteins. Results expressed as spot-forming units per million cells (SFU/106). The dashed line represents a positivity threshold of the mean + 2SD of the background response. Data shown with median and interquartile range. Median fold-change from pre- to post-infection samples is displayed. Statistical comparisons of paired pre- and post-infection samples done with Wilcoxon signed-rank test, and between post-infection levels in previously-infected and SARS-CoV-2 naive individuals using the Mann-Whitney U test. P values are displayed where <0.05. Responses were evaluated in 37 SARS-CoV-2-naive and 32 previously-infected individuals for whom samples were available.

### Evaluation of the heterogeneity in SARS-CoV-2 immune responses before and after omicron infection

A principal component analysis was performed to view the heterogeneity in SARS-CoV-2 immunity when integrating the pre- and post-omicron mucosal and blood immune responses in the 94 individuals included in the study (Figure 5). 44.6% of the variance observed in the first two dimensions (PC1 and PC2) could be explained by the measured immunological parameters. Omicron infection was a major driver for separation in the data, although there was considerable variability in the impact of omicron across individuals (Figure 5A). Prior infection status also had a major effect on separation in the data, although, again, considerable overlap was present across individuals (Figure 5B). Two distinct patterns of immunity were observed, driven either by plasma binding and NAb responses (Figure 5C, *lower right quadrant*, highly correlated with dimension 1), or T cell responses (Figure 5C, *upper right quadrant*, highly correlated with dimension 2; Figure 5D). Plasma antibody and blood T cell responses were the most important factors in immunophenotypic variability, with limited impact of variables such as age or time between 3rd vaccine dose to omicron infection (Figure 5E).

**Figure 5.**
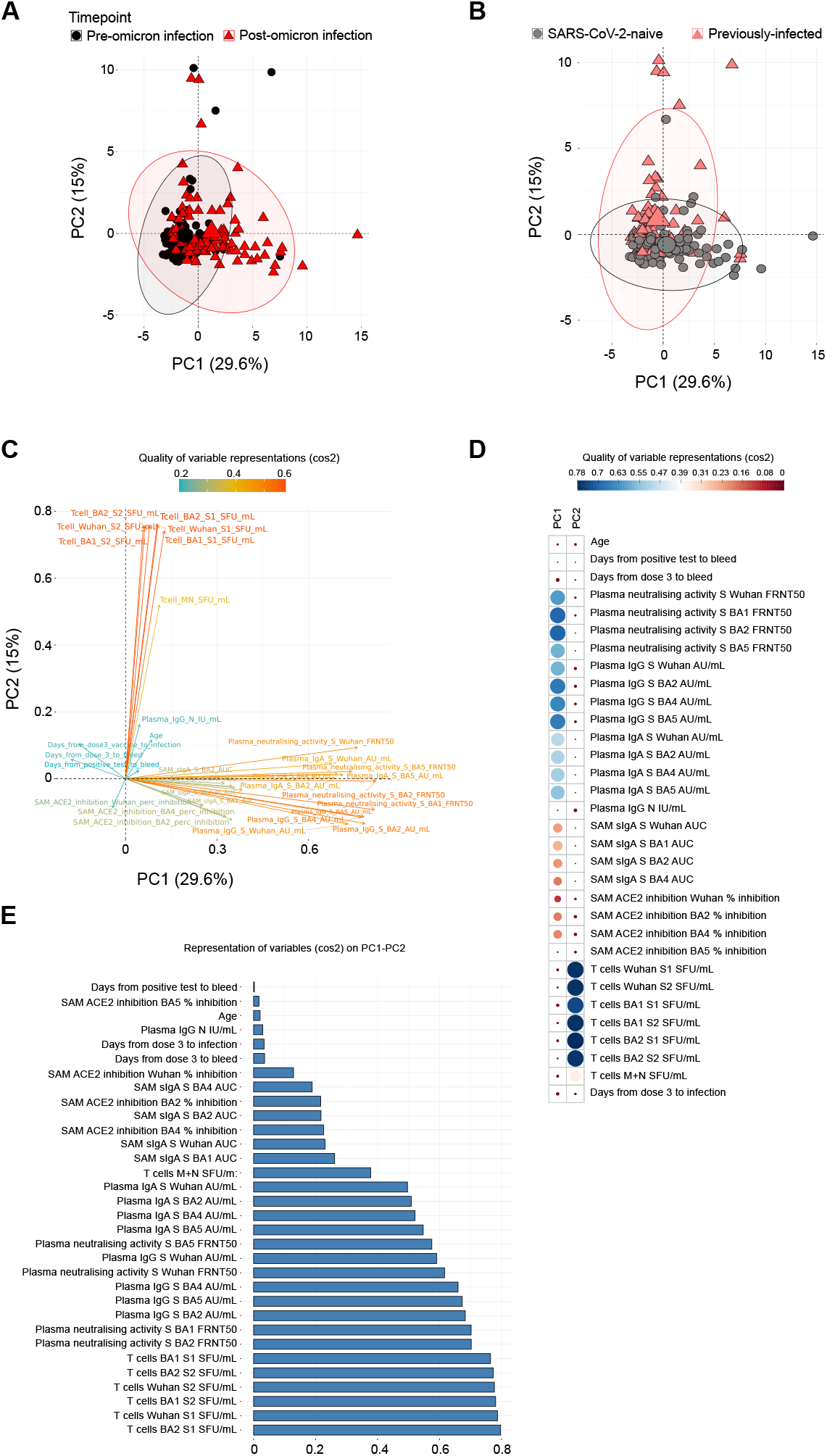
Principal Component Analysis (PCA) of immune responses before and after SARS-CoV-2 omicron infection. PCA plot representing integrated immunological data, representing components 1 (PC1) and 2 (PC2) annotated by samples from pre-and post-omicron infection (A) and from SARS-CoV-2 naive and previously-infected individuals (B). Percentages indicate the variance explained by PC1 and PC2. (C) Variable correlation plot, where positively correlated immune responses are grouped together and negatively correlated variables are found in opposite quadrants. The colour indicates the quality of representation of the variable on the principal component (cos2), with higher cos2 equating to greater representation. (D) Quality of variable representations coloured by cos2 and contributions of variables to PC1 and PC2 (size of circle, larger circle = greater contribution). (E) All variables included in the PCA, ordered by degree of representation on PC1 and PC2 (cos2).

### Impact of omicron infection on spike- and non-spike epitope-specific CD8+ T cells

Given the differences in T cell responses observed between SARS-CoV-2 naive and previously infected healthcare workers pre- and post-omicron infection, as well as distinct patterns of response to spike and non-spike proteins, detailed phenotypic characterization of SARS-CoV-2 epitope-specific CD8+ T cells was performed using major histocompatibility complex (MHC) class I-peptide multimer staining and multiparameter flow cytometry. Immunodominant spike (A*01:01, A*02:01, A*03:01, B*57:01) and non-spike (A*01:01 NSP3, B*07:02 nucleocapsid) epitope-specific CD8+ T cells were characterized, along with cytomegalovirus (CMV)- and Epstein-Barr virus (EBV)-specific CD8+ populations as controls. In keeping with the IFN-***γ*** ELISpot responses, spike-specific CD8+ T cells increased in magnitude following omicron infection in SARS-CoV-2 naive (p=0.02), but not previously-infected individuals, whereas non-spike populations increased significantly in both groups (p=0.002 and p=0.001; Figures 6A and 6B). No changes in EBV- or CMV-specific CD8+ T cells were seen.

**Figure 6.**
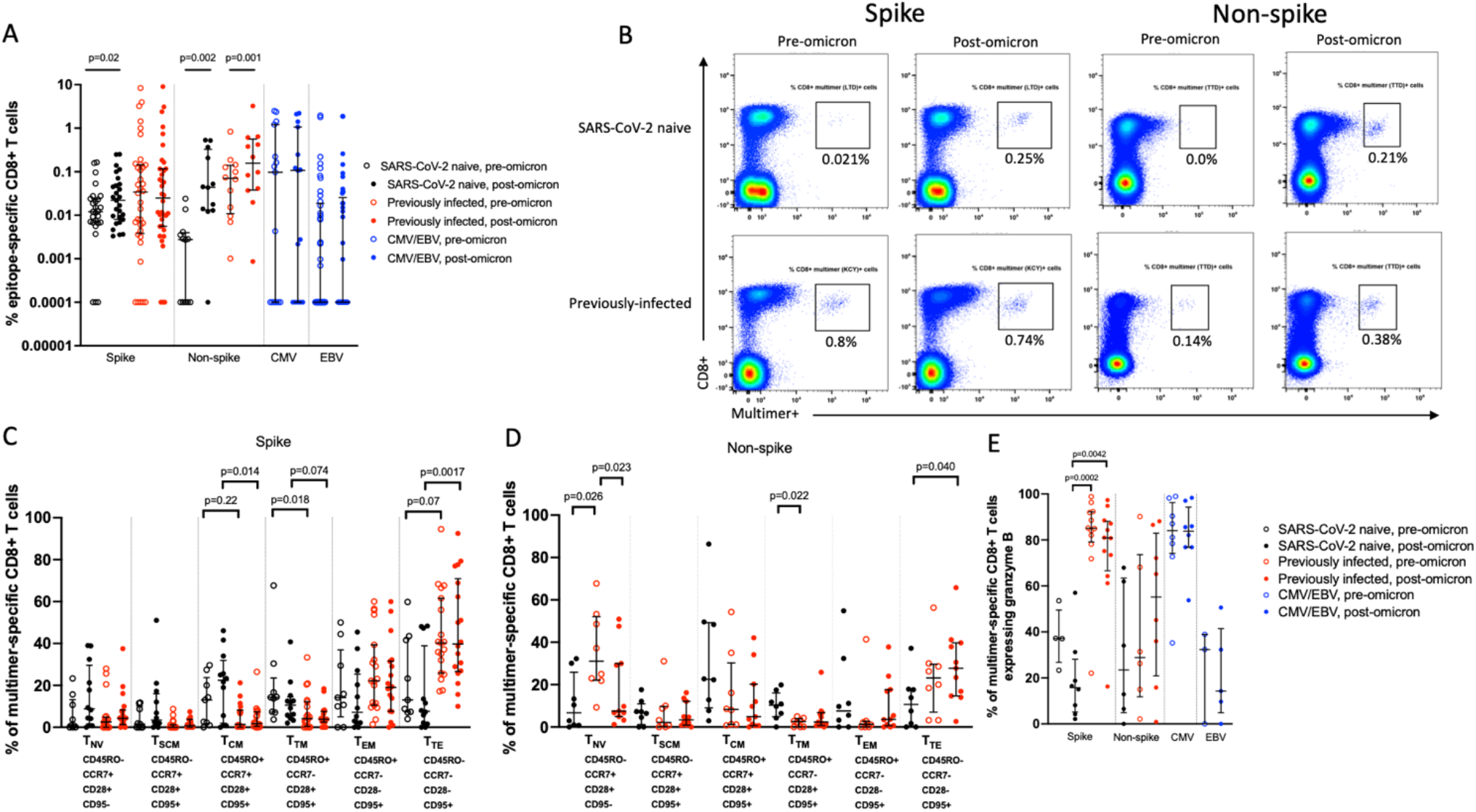
Epitope-specific CD8+ T cell magnitude and phenotype before and after omicron infection. (A) Magnitude of multimer-specific CD8+ T cells expressed as a % of total CD8+ cells following mRNA vaccine dose 3 (pre-omicron) and after omicron infection (pooled data from Sheffield and Newcastle). Shown are 63 spike-specific multimer populations from 45 individuals (23 naive, 22 previously-infected), 24 non-spike populations from 20 individuals (10 naive, 10 previously-infected), 15 CMV-specific populations from 15 individuals, and 41 EBV-specific populations from 32 individuals. A value of 0.0001% is assigned to negative samples for the purpose of display on a log10 axis (B) Representative flow cytometry plots showing multimer-specific spike- and non-spike CD8+ populations from naive and previously-infected individuals before and after omicron infection (C) Memory phenotypes of spike-specific CD8+ T cells in 11 SARS-CoV-2 previously-naive and 14 previously-infected individuals (pooled data from Sheffield and Newcastle). (D) Memory phenotypes of non-spike CD8+ T cells in 7 SARS-CoV-2 previously-naive and 10 previously-infected individuals (pooled data from Sheffield and Newcastle) (E) Granzyme B expression in epitope-specific CD8+ T cells (data from Sheffield). Shown are spike-specific populations from 8 naive and 12 previously-infected individuals, non-spike populations from 6 naive and 8 previously infected individuals, and CMV- and EBV-specific populations from 8 and 5 individuals respectively. Data shown with median and interquartile range. Statistical comparisons of paired pre- and post-infection samples where available from the same participants were done with Wilcoxon signed-rank test, and between unpaired comparisons using the Kruskal-Wallis test and Dunn’s post-hoc test for multiple pairwise comparisons. P values are >0.05 unless displayed. T_NV_ = naive T cells, T_SCM_ = stem cell memory T cells, T_CM_ = central memory T cells, T_TM_ = transitional memory T cells, T_EM_ = effector memory T cells, T_TE_ = terminal effector T cells.

Epitope-specific T cells were classified into different memory phenotypes as previously described using combinations of CD45RO, CCR7, CD28 and CD95 expression into naive (T_NV_), stem cell memory (T_SCM_), central memory (T_CM_), transitional memory (T_TM_), effector memory (T_EM_), and terminal effector (T_TE_) T cells (15). After the 3rd mRNA vaccine dose, but prior to omicron infection, spike-specific CD8+ T cells in previously-infected individuals displayed lower T_NV_ phenotypes (p=0.0027) and higher T_EM_ phenotypes (p=0.016) than non-spike CD8+ T cells (Figure S3). Visualization of cell clusters using Uniform Manifold Approximation and Projection (UMAP) plots showed distinct populations of CMV and EBV-specific cells, with some separation of SARS-CoV-2 specific CD8+ T cell clusters, driven by differences in expression levels of several markers (Figure S4).

The phenotype of spike-specific CD8+ T cells did not change significantly from before to after omicron infection in either previously-naive or -infected individuals (Figures 6C, S5 and S6). Nevertheless, key differences were seen between the two groups, with spike-specific CD8+ populations in previously-infected individuals displaying fewer T_CM_ and T_TM_ T cells and a more terminal effector phenotype than naive vaccinees, even after omicron infection (p=0.0017, Figure 6C). Non-spike CD8+ populations in previously-infected individuals consisted of fewer cells with a T_NV_ phenotype after omicron infection (p=0.023, Figure 6D). These boosted CD8+ T cells were also more likely to display a terminal effector phenotype than newly primed non-spike populations in the previously-naive group (p=0.040, Figure 6D). Expression of the cytolytic enzyme granzyme B in spike-specific CD8+ T cells did not change significantly after omicron infection, however, expression levels were significantly higher in previously-infected individuals at both timepoints (p=0.0002 and p=0.0042, Figure 6E), to a level similar to that seen in CMV-specific T cells.

## Discussion

In countries with high SARS-CoV-2 vaccination rates, the emergence of omicron lineage viruses with considerable immune escape from vaccine-induced NAbs has led to high levels of infection followed by hybrid immunity. The nature of this post-omicron immunity has been questioned in two main ways by recent findings (12,13). Firstly, it has been suggested that the relatively attenuated nature of omicron infections (11) results in poor induction of immune responses, and secondly that ‘imprinting’ from previous SARS-CoV-2 infection (e.g. with ancestral or alpha viruses) leads to profound immune-dampening; with both scenarios precluding the generation of omicron-specific immunity able to protect against future omicron infections. These concerns were raised on the basis of data focused primarily on circulating NAbs, mediated by spike-specific responses. We have taken a broader approach and characterized mucosal and circulating immunity to spike and non-spike antigenic targets. While we replicate some of the previously reported NAb data, we also demonstrate that omicron infections result in significant increases in mucosal antibodies and circulating T cells, with the dominant antigenic targets of some responses dependent on prior SARS-CoV-2 infection history.

We and others have repeatedly shown that individuals with a history of SARS-CoV-2 prior to their 1st mRNA vaccine have higher NAb titres after each SARS-CoV-2 vaccine dose when compared to infection-naive individuals (8–10). This advantage is eliminated immediately after omicron infection in triple-vaccinated adults, with omicron-specific NAbs getting a greater boost in previously-naive healthcare workers. This unexpected finding in triple-vaccinated individuals was first reported by Reynolds *et al*. (13), who noted that post-omicron NAb titres in 11 previously-naive healthcare workers were significantly higher than in 6 individuals with a history of infection with pre-alpha SARS-CoV-2 viruses. Our larger cohort confirms this finding, but also demonstrates a great deal more heterogeneity, with 70% of previously-infected individuals increasing omicron-specific NAb titres in response to BA.1 or BA.2 infection. A study of 56 triple-vaccinated healthcare workers in Sweden also demonstrated that post-omicron IgG and NAb levels were higher in previously-naive individuals compared to those with a history of prior SARS-CoV-2 infection infections (12). Importantly, omicron-specific antibody immunity was still higher in both groups compared to healthcare workers who did *not* have breakthrough omicron. Several other studies have confirmed that omicron infections in previously-naive triple-vaccinated adults increase omicron-specific NAb activity (16), primarily through expansion of memory B cells against epitopes that are conserved across variants.

The mechanism behind the relatively poor induction of circulating NAbs by omicron infection in triple-vaccinated previously-infected individuals remains unexplained. Imprinting or original antigenic sin (OAS) from prior infections has been suggested as a possible explanation (13). The concept of OAS was first described for influenza antibodies (17), with imprinting from prior influenza infections on immune responses following subsequent influenza infection and vaccination noted in more recent work (18–20). OAS from antibodies generated by prior OC43/HKU1 seasonal coronavirus infections occurs; and is seen more potently following SARS-CoV-2 infection than after the 1st mRNA vaccine dose, with SARS-CoV-2 infection driving a boost of lower affinity antibodies (21). Potentially beneficial imprinting has also been described; omicron breakthrough infections in infection-naive vaccinated individuals induce antibodies from B cells that cross-react with receptor-binding domains from multiple variants and provide a greater breadth of protective immunity whereas antibodies following primary omicron infections in infection-naive unvaccinated individuals are of much narrower specificity (16). Of note, we did not observe a boosting of NAbs against an ancestral strain of SARS-CoV-2 at the expense of generating omicron-specific immunity. Importantly, even individuals with prior infection had a much greater increase in BA.1/BA.2/BA.5 NAb activity than to the ancestral strain. While it is likely that conserved epitopes between ancestral and omicron viruses would have been preferentially boosted over omicron-specific epitopes, it is unlikely that OAS alone fully explains the attenuated response in previously-infected individuals we observed.

Another hypothesis is that the higher omicron-specific immunity after the 3rd mRNA vaccine dose seen in those with prior SARS-CoV-2 infection may lead to lower viral loads and therefore reduced antigenic exposure during omicron infection, resulting in muted immune induction. While Blom *et al* (12) did find a correlation between cycle threshold (Ct) values and NAb levels, nadir Ct values (i.e. highest viral loads) were no different between naive and previously-infected healthcare workers. The amount of virus in previously-infected individuals in our study was certainly sufficient to generate significant increases in non-spike antibodies and T cells; however, this does not rule out the possibility that antigenic load was lower in this group, impacting the post-omicron immunity observed. Finally, it is possible that differences in spike-specific B cell phenotype could underlie the variability in NAb responsiveness to omicron infection. A link between the frequency of CD27lo B cells and altered B cell receptor (BCR) signaling with the response to 3rd mRNA vaccine dose has been reported (22). Although in that particular study recent infection prior to vaccination drove a B cell phenotype that led to muted vaccine response, future work should explore whether molecular mechanisms that impair BCR signaling are more prevalent in previously-infected individuals after a 3rd mRNA vaccine dose than in naive triple-vaccinated adults.

We found that spike-specific mucosal antibodies were low after the 3rd mRNA vaccine dose and similar between SARS-CoV-2 naive and previously-infected individuals. We and others have shown that mucosal IgA is induced by mRNA vaccines in those with a history of prior SARS-CoV-2 (23,24), so this finding was unexpected, and is perhaps explained by our cohort being enriched for previously-infected healthcare workers with low mucosal immunity and therefore at risk of breakthrough infection. We have also previously noted a waning of mucosal IgA over 9 months after SARS-CoV-2 infection (25) and the time to first infection in our previously-infected cohort was much longer. Nevertheless, omicron infection induced mucosal SARS-CoV-2-specific secretory IgA and omicron-specific NAb responses, regardless of prior SARS-CoV-2 infection history. Mucosal immunity is likely to be important both for protection from infection and transmission-blocking immunity, yet is poorly induced by currently licensed parenteral vaccines. Our findings and those from others (16,26) suggest that omicron breakthrough infections in vaccinated individuals may contribute to filling this immunity gap.

We demonstrate that omicron infection increased spike-specific T cell responses in previously-naive individuals, and non-spike-specific T cells regardless of prior SARS-CoV-2 history, with membrane and nucleocapsid-specific T cells boosted to a greater degree in previously-infected healthcare workers. These findings are in contrast to those in Blom *et al*. (12), who found no increase in spike-specific responses after omicron infection using the Oxford Immunotech (OI) T-spot assay, and similar M- and N-specific T cell responses in both previously-infected and -naive individuals post-omicron. These differences may be explained by the assays used, as we have previously reported higher magnitude responses in our PITCH ELISpot assay when compared to the OI assay, which is not marketed as a quantitative test (27). In our study, despite not increasing spike-specific T cells following omicron infection, previously infected individuals still maintained higher S1-specific responses than their previously-naive counterparts.

Detailed characterisation of immunodominant CD8+ spike epitope-specific cells in previously-infected individuals also demonstrated a more terminally differentiated, highly cytotoxic phenotype. The high granzyme B content in these T cells was strikingly similar to that seen in CMV-specific CD8+ T cells, shown previously to have a late-differentiated phenotype with high cytotoxic potential distinct from other anti-viral CD8+ T-cells (28). Our findings suggest that an infection prime followed by 3 mRNA vaccine doses results in maximally induced spike-specific T cell responses, with limited potential for further boosting, at least in the short term. Interestingly, the granzyme B content of spike-specific CD8+ T cells in previously-naive individuals *after* omicron infection was much lower than in previously-infected individuals *prior* to omicron. As both these scenarios represent immunity after four spike exposures, the order of exposures may matter in the generation of hybrid immunity with respect to the cytotoxic potential of CD8+ T cells. T cell responses to non-spike targets increased significantly in all individuals following omicron infection, with those with prior SARS-CoV-2 having higher post-omicron levels. A similar advantage in nucleocapsid-specific IgG was observed in previously-infected individuals following omicron infection. While these antibodies would not be expected to contribute to NAb activity, a protective role through antibody-dependent cellular cytotoxicity or other non-neutralizing effector functions may exist (29,30).

The key question is whether omicron infections in vaccinated individuals confer a degree of protection against subsequent omicron lineage viruses. Real-world effectiveness data are aligned with our findings that omicron infections induce or boost diverse immune responses in vaccinated individuals. In Portugal, where 98% of the study population had completed a primary 2-dose vaccine course prior to 2022, and mRNA vaccine booster coverage was 82% at the start of the BA.5 wave, the protective efficacy of BA.1/BA.2 breakthrough infections against subsequent BA.5 infection was 75.3% compared to infection-naive vaccinees, whereas previously efficacy following breakthrough infections with other variants was lower (e.g. 54.8% following alpha, 61.3% following delta) (31). Similarly, prior omicron infection in triple-vaccinated individuals was found to be 93.6% protective against subsequent BA.5 infection in a Danish cohort study (32).

Our study has several limitations which need to be considered and may affect the generalisability of our findings. Most importantly, we selected previously-infected individuals who had breakthrough omicron infections within a few months of a 3rd mRNA vaccine dose, and who therefore may not be immunologically similar to those who remained protected for longer. Real-world protection from symptomatic BA.1/BA.2 infection is greater in those with infection followed by three mRNA vaccine doses compared to SARS-CoV-2 naive triple-vaccinated individuals (33), therefore we likely enriched for previously-infected individuals with lower protective immunity. In most experiments, we also deliberately considered responses to whole proteins rather than specific epitopes to capture the breadth of immunity generated through SARS-CoV-2 infection. It is likely that individual epitope-specific differences may exist between groups, including those affected by mutations in omicron lineage viruses.

Although vaccination remains the safest way to acquire SARS-CoV-2 immunity, accumulating data suggest that infection-induced immunity in SARS-CoV-2 vaccinees can provide enhanced protection. For example, protection against BA.4/5 infection is estimated to be more durable in UK adults with a delta/omicron breakthrough infection after two vaccine doses, compared with triple-vaccinated adults with no history of infection (34). To date, there is epidemiological evidence that hybrid immunity gives good protection against BA.4/5, for example protection against hospitalization with BA.4/5 was greater than 90% for at least 6-8 months in a Quebec study of adults over 60 years of age who had received at least two vaccine doses and one infection at any time (35). As highly vaccinated populations experience increasing numbers of SARS-CoV-2 infections, the nature of hybrid immunity in individuals becomes more complex and heterogeneous. We have demonstrated that immune components not induced by currently available vaccines such as mucosal and non-spike responses are enhanced by viral infection. These may play a critical role in accumulating protective immunity to SARS-CoV-2, but should also be prioritised as targets to broaden the responses generated by the next generation of vaccines.

## Materials and Methods

### Participant recruitment

The PITCH (Protective Immunity from T cells to Covid-19 in Health workers) study is a prospective cohort study of 2149 healthcare workers (HCWs) recruited at five sites in the UK (University Hospitals Birmingham NHS Foundation Trust, Liverpool University Hospitals NHS Foundation Trust, Newcastle upon Tyne Hospitals NHS Foundation Trust, Oxford University Hospitals NHS Foundation Trust, and Sheffield Teaching Hospitals NHS Foundation Trust). Eligible participants were adults aged 18 years and over, currently working as health-care workers, including allied support and laboratory staff. Individuals were recruited by word of mouth, hospital email communications, from hospital-based staff SARS-CoV-2 screening programmes, as well as enrolling through the wider SIREN study, of which PITCH is a substudy. The SIREN study is registered with ISRCTN (Trial ID:252 ISRCTN11041050), and was approved by the Berkshire Research Ethics Committee, Health Research 250 Authority (IRAS ID 284460, REC reference 20/SC/0230), with PITCH recognized as a sub-study on 2 December 2020. Some participants were recruited under other protocol-aligned REC-approved studies; in Liverpool some participants were recruited under the ‘‘Human immune responses to acute virus infections’’ Study (16/NW/0170), approved by North West - Liverpool Central Research Ethics Committee on 8 March 2016, and amended on 14th September 2020 and 4th May 2021. In Oxford, participants were recruited under the GI Biobank Study 16/YH/0247, approved by the research ethics committee (REC) at Yorkshire & The Humber - Sheffield Research Ethics Committee on 29 July 2016, which has been amended for this purpose on 8 June 2020. In Sheffield, participants were recruited under the Observational Biobanking study STHObs (18/YH/0441), which was amended for this study on 10 September 2020. All procedures were conducted in accordance with the principles of the Declaration of Helsinki (2008), and the International Conference on Harmonization Good Clinical Practice guidelines. All participants enrolled provided written informed consent.

Participants were defined as either SARS-CoV-2 naive or previously infected at the time of enrolment in the PITCH study based on documented PCR and/or serology from local NHS trusts, or from MSD analysis of S and N plasma IgG levels of PITCH samples (8).

Patients were sampled 28 days following their third vaccine dose, and 28 days following documented SARS-CoV-2 infection. At each visit synthetic absorption matrix (SAM) strips were collected to obtain nasal lining fluid, and heparinised whole blood collected, which was separated via density gradient centrifugation into plasma and peripheral blood mononuclear cells (PMBCs). Nasal lining fluid was sampled by inserting the SAM strip into the nostril and holding against the mucosa for 1 minute via light finger pressure on the outside of the nose.

### NAb responses

The neutralizing ability of plasma was measured using a Focus Reduction Neutralization Test (FRNT). Briefly, serially diluted plasma was mixed with live SARS-CoV-2 virus of an ancestral strain (Australia/VIC01/2020), BA.1, BA.2, or BA.5, then incubated at 37°C for 1 hour. The plasma-virus mixture was then transferred to 96-well, cell culture-treated, flat-bottom microplates with confluent monolayers of Vero cells in duplicate and incubated at 37°C for further 2 hours. After incubation, 1.5% carboxymethyl cellulose (CMC) overlay medium diluted in Dulbecco’s Modified Eagle Medium (Gibco, 31966047) with 1% FBS (DMEM1) was added into each well. A focus forming assay was performed after cells had been incubated with virus at 37°C for 24 hours. Vero cells were fixed with 4% Formaldehyde (Sigma, F8775), permeabilized with 2% Triton-X (Sigma, T9284), then stained with human anti-N mAb (mAb206) followed by peroxidase-conjugated goat anti-human IgG (Sigma, A0170). After staining, the foci were visualised by adding TrueBlue Peroxidase Substrate (SeraCare, 5510-0030), and approximately 100 foci were observed in the wells without plasma. Infection plates were counted on the AID EliSpot reader using AID ELISpot software. The percentage of focus reduction was calculated for each plasma and FRNT50, the reciprocal dilution of plasma required to neutralize 50% of input virus, was determined using the probit program from the SPSS package.

### Spike-specific IgG and IgA binding antibody responses

SARS-CoV-2-specific IgG and IgA binding antibody levels in plasma were assessed using the multiplex Mesoscale Discovery (MSD) platform V-PLEX SARS-CoV-2 Panel 27 IgG (Meso Scale Diagnostics, K15606U) and V-PLEX SARS-CoV-2 Panel 27 IgA (Meso Scale Diagnostics, K15608U) kits. Assays were performed as per the manufacturer recommended protocol. Plates pre-coated with SARS-CoV-2 spike antigen spots (including Wuhan-Hu-1, and omicron sublineages BA.2, and BA.5) were blocked with 150 µL Blocker A solution for 30 minutes at room temperature (RT), shaking at 800 rpm. No BA.1 sublineage protein was present on kit 27 plates. Plates were washed, and 50 µL/well plasma samples diluted to 1:40000 in Diluent 100 were loaded in duplicate and incubated for 2 hours at RT, shaking at 800 rpm. Plates were washed, and 50 µL/well of 1X detection antibody solution was added to each well and incubated for 1 hour, shaking at 800 rpm. Plates were washed and 150 µL MSD GOLD Read Buffer B was added to wells, before reading immediately with a MESO® SECTOR S 600 instrument.

### Nucleocapsid IgG antibody detection

Nucleocapsid IgG was assessed using an ELISA as previously described (36). High-binding 96-well ELISA plates (Immulon 4HBX; Thermo Scientific, 6405) were coated overnight at 4°C with 50 µL/well full-length untagged nucleocapsid protein produced in *E. coli* (Uniprot ID P0DTC9 (NCAP_SARS2), diluted to 2 µg/mL in 7.4 pH phosphate buffered saline (PBS). Plates were washed with 0.05% PBS-Tween, then blocked for one hour with 200 µL/well 0.5% casein buffer. Plasma samples were diluted to 1:200, and 100 µL loaded in duplicate wells. Plates were incubated for 1 hour at RT, then washed and loaded with 100 µL/well of goat anti-human IgG-HRP conjugate (Invitrogen, 62-8420) at 1:500. Plates were incubated for 2 hours at RT, then washed and developed for 10 minutes with 100 µL/well TMB substrate (KPL, 5120-0074) and stopped with 100 µL/well HCl Stop solution (KPL, 5150-0021). Absorbance at 450 nm (A450) was read immediately with a HIDEX sense luminometer.

To allow quantification of antibody concentration, we included a 12-step standard curve consisting of sera pooled from convalescent SARS-CoV-2-confirmed patients, calibrated to the WHO International Standard for anti-SARS-CoV-2 immunoglobulin (NIBSC, 20/136), with results reported in binding antibody units/mL (BAU/mL). Samples that were considered negative based on previously determined thresholds (36) were assigned a value of 1.04 BAU/mL, which was half the value of the lowest point on the standard curve.

### Secretory IgA

Levels of spike- and nucleocapsid-specific dimeric secretory IgA (sIgA) present in nasal lining fluid were assessed in an ELISA using a primary antibody targeting the human secretory component. Levels of total sIgA were also assessed to allow normalisation of SARS-CoV-2-specific sIgA results.

To detect SARS-CoV-2 antibodies, high binding 96-well ELISA plates (Immulon 4HBX; Thermo Scientific, 6405) were coated overnight at 4°C with 50 µl/well of SARS-CoV-2 spike proteins representing the SARS-CoV-2 D614G (Sino Biological 40589-V08H8), omicron BA.1 (40589-V08H26), BA.2 (40589-V08H28) or BA.4 (40589-V08H32) viruses, diluted to 1 µg/mL in 7.4 pH PBS, or full-length untagged nucleocapsid protein produced in *E. coli* (Uniprot ID P0DTC9 (NCAP_SARS2), diluted to 2 µg/m in 7.4 pH PBS. For total sIgA ELISAs, plates were coated overnight with goat anti-human kappa and lambda light chain antibodies (Southern Biotech, 2060-01, 2070-01) each diluted to 2 µg/mL in Dulbecco’s phosphate buffered saline (DPBS). Plates were washed with 0.05% PBS-Tween, and blocked for one hour with 200 µL/well 1% casein buffer.

For detection of SARS-CoV-2-specific sIgA, samples were tested in duplicate in a 5-well dilution series proceeding in 2x steps from 1:10 to 1:160. Plates were loaded with 50 µL/well of sample and incubated overnight at 4°C. A curve was generated for each sample by plotting A_450_ against a dilution coefficient. This was used to calculate an area under the curve (AUC) value for each sample. To detect total sIgA, nasal lining fluid was tested in duplicate wells at 1:4000. For quantification purposes a standard curve of human IgA from colostrum (Sigma, I2636) was included in a 12-well dilution series proceeding in 2x steps from an initial dilution of 1 µg/mL. Plates were incubated for 1 hour at 36°C. Following sample incubation, plates (both specific and total) were washed and loaded with 100 µL/well mouse anti-human secretory component antibody (Calbiochem, HP6141) at 1 µg/mL for two hours at RT. Plates were washed and loaded with goat anti-mouse IgG-HRP conjugate (Invitrogen, 31439) at 1:500 for 1 hour at RT, then washed and developed for 5 mins with 50 µL/well TMB substrate (KPL, 5120-0074) before addition of 50 µL/well HCl stop solution (KPL, 5150-0021). A450 was read immediately with a HIDEX sense luminometer. SARS-CoV-2 specific sIgA AUC values were normalized to total sIgA from the same sample.

### Surrogate neutralizing activity in nasal lining fluid

SARS-CoV-2-specific surrogate neutralizing ability of nasal lining fluid was assessed using the MSD V-PLEX SARS-CoV-2 Panel 27 ACE2 kit (Meso Scale Diagnostics, K15609U), which assesses the ability of samples to prevent binding of ACE2 to SARS-CoV-2 spike antigens, including Wuhan-Hu-1, and omicron sublineages BA.2, and BA.5. SAM strips were eluted into a buffer of PBS, 1% BSA, and protease inhibitor cocktail I (Calbiochem, 539131). Plates pre-coated with SARS-CoV-2 antigen spots were blocked with 150 µL Blocker A solution for 30 minutes at RT, shaking at 800 rpm. Plates were washed, and eluted nasal lining fluid samples were loaded neat at 25 µL/well in duplicate and incubated for 1 hour at RT, shaking at 800 rpm. After incubation, 25 µL of 1X SULFO-TAG Human ACE2 Protein solution was added to wells without washing or aspiration of sample, and incubated for 1 hour at RT, shaking at 800 rpm. Plates were washed, and 150 µL/well MSD GOLD Read Buffer B added, before reading immediately with a MESO® SECTOR S 600 instrument. The neutralizing activity of samples was expressed as the percentage of ACE2 inhibition compared to a condition with no nasal lining fluid on the same plate.

### IFN-γ ELISpot assays

IFN-***γ*** ELISpot assays were performed on cryopreserved peripheral blood mononuclear cells (PBMCs) using the ELISpot Flex Human IFN-***γ*** kit (Mabtech, 3420-2A). Assays were performed in Sheffield, Oxford, and Newcastle, using the same previously harmonised and published protocol (8). Overlapping peptide pools (18-mer, with 10 amino acid overlap) representing the S1 and S2 subunits of SARS-CoV-2 Wuhan-hu-1, omicron BA.1 and omicron BA.2 spike proteins, pooled ancestral Wuhan-hu-1 membrane and nucleocapsid proteins, and 6 non-structural proteins (NSP1+2, NSP3b+c, NSP4-6, NSP7-11, NSP12, and NSP13+14) were used to stimulate PBMCs at 2 µg/mL. Pools of CMV, EBV and influenza peptides (CEF) and phytohemagglutinin (PHA) were also included as positive controls, as well as a cell only condition. Sterile 96-well plates with 0.45 µm PVDF membrane were coated with 50 µL/well of anti-IFN-***γ*** coating antibody diluted to 10 µg/mL in sterile PBS, and incubated overnight at 4°C. Plates were washed with sterile PBS and blocked with 200 µL/well R10 (RPMI + 10% FBS + 1% pen/strep). After at least 2 hours incubation at 37°C, wells were emptied, and 200,000 cells diluted in 50 µL R10 were added to each well. 50 µL of peptide pools were added to wells, and plates were incubated for 18-24 hours at 37°C, washed and 100 µL/well of biotinylated secondary antibody diluted to 1µg/mL in PBS 0.5% BSA loaded before incubation for 2-4 hours at RT. Plates were washed and 100 µL/well of streptavidin-ALP conjugate diluted in PBS to 1 mg/mL was added to wells and incubated for 40 minutes at RT. Plates were developed using the AP conjugate substrate kit (Biorad, 170-6432). Plates were washed and 100 µL of detection solution added to wells and incubated for 15 minutes at RT. Plates were washed with tap water and left to dry before reading on the AID EliSpot reader. Spots were counted using AID ELISpot software, duplicate wells were averaged and cell only value subtracted, and virus-specific responses were expressed as spot-forming units (SFUs)/10^6^ cells. A cut-off for positivity was determined by taking the mean + 2SD of the SFU/10^6^ cell value of all cell only control wells. Negative results were assigned a value of 1 SFU/10^6^.

### DNA extraction and HLA typing

DNA was isolated from cryopreserved whole blood samples using the DNeasy® Blood and Tissue kit (QIAGEN, 69504) as per the manufacturer’s protocol. Newcastle based participant HLA typing for Class I (HLA-A; B; C) was performed by MC Diagnostics Limited (Wales, United Kingdom). Sheffield participant HLA Class I typing was performed at The University of Oxford by amplifying Exons 2 and 3 for each locus using in-house sequence-specific primers and sequenced on an Applied Biosystems AB3730 instrument. Analysis was done by comparing heterozygote traces to known sequences on the IMGT-HLA database to 4-digit resolution.

### Multi-parameter flow cytometry characterising antigen-specific CD8+ T cells

Cryopreserved PBMCs were rapidly thawed and rested for 2-4 hours in R10 media (RPMI + 10% FBS + 1% pen/strep) at 37°C. After resting and prior to staining, cells were washed in PBS and divided into FACS tubes at 2-3 × 10^6^ cells per sample. Firstly, samples and HLA mismatched negative control donors were stained with PE conjugated SARS-CoV-2 spike or non-spike specific MHC pentamer/dextramers (PE-HLA-A*03:01 KCYGVSPTK S_378,_ ProImmune, peptide code 4443, PE-HLA-A*02:01 YLQPRTFLL S_269_, PE-HLA-A*01:01 LTDEMIAQY S_865_, PE-HLA-B*57:01 GTITSGWTF S_879_, PE-HLA-A*01:01 TTDPSSFLGRY RP_1637_, PE-HLA-B*07:02 SPRWYFYYL NCP_105_, Table S3.) and APC conjugated EBV and CMV pentamer/dextramers (APC-HLA-A*02:01 GLCTLVAML BMLF-1_259_, APC-HLA-B*07:02-RPPIFIRRL EBNA-3A_247_, APC-HLA-A*03:01 RLRAEQVK EBNA-3A_603_ or APC-HLA-A*02:01 NLVPMVATV CMV pp65_495_, Table S3.) for 15 minutes at 37°C. Surrogate CD3 conjugates for PE and APC were used as reference controls for unmixing either by staining cells (Newcastle) or beads (Sheffield). After pentamer/dextramer staining, samples were washed in PBS and re-suspended in the residual volume (∼50μl). Samples were stained with Live/Dead Zombie NIR (Biolegend, 423106) for 10 minutes at RT, after which and without washing, the surface antibody cocktail diluted in Brilliant Stain Buffer (Becton Dickinson, 563794) was added for a further 20 minutes. Single stain reference controls were stained as per Tables S1 and S2 on 2-5 × 10^5^ cells/mL or UltraComp eBeads(tm) (ThermoFisher, 01-2222-42). The Live/Dead Zombie NIR reference control was prepared by staining a 50/50 mixture of live and heat inactivated (56°C, 7 minutes) PBMCs. Following the extracellular staining, samples prepared in Sheffield were also stained for granzyme B using the BD Cytofix/Cytoperm(tm) Kit (Cat. No. 554714) according to the manufacturer’s instructions. Samples and reference controls in Newcastle were washed in PBS after extracellular staining and fixed for 20 minutes in 2% formaldehyde at RT. After fixation, samples were washed in FACS buffer and re-suspended in an appropriate volume for acquisition on a CyTEK AURORA 3L (Sheffield) or 5L (Newcastle) system. Data were unmixed using a combination of bead and cell reference controls, as outlined in Tables S1 and S2, and post-unmixing compensation applied where required.

### Data Analysis

Statistical analysis was performed using Graphpad Prism 9.4.1 for Windows. Comparisons between continuous data from SARS-CoV-2-naive and previously infected groups were performed using Mann Whitney tests or Kruskal Wallis tests with Dunn’s post-hoc test for multiple comparisons, and categorical data compared with the Fisher’s exact test. Pairwise comparisons within groups were performed using Wilcoxon matched-pairs signed-rank tests. Principal component analysis (PCA) was performed using SIMON software version 0.2.1 (https://genular.org) (37). Before PCA was performed, the data was pre-processed (center/scale), missing values were median imputed and variables with fewer than 5 unique values were removed. The *sex, site, previous infection status* and *timepoints* were used as grouping variables, and thus were not included in the analysis. The quality of individuals and variable representations (cos2), variable correlations and contributions (percentage) of top 10 variables from the first two principal components (PC1 and PC2) were calculated.

Gating analysis of flow cytometry data was performed in SpectroFlo® (version 3.0.0). Data were gated for live, CD3+CD8+ pentamer/dextramer+ populations from twelve batches across two different sites (Sheffield, Newcastle). HLA mismatched pentamer/dextramer stains were used to determine gating for positive events (Figure S6). A pentamer/dextramer event rate of ≥20 (2-dimensional gating, combined data from both sites) or ≥0.01% of the total CD8+ population (multi-dimensional clustering, data from sites analysed separately) was used as a threshold to take forward populations for further analysis.

Multi-dimensional analysis of flow cytometry data was undertaken in R using the packages readxl, CATALYST, cowplot, flowCore, scater, SingleCellExperiment, openxlsx and ggpubr. FCS data from multimer-positive gates were transformed using a cofactor of 150 and FlowSOM clustering applied to all channel markers present on pentamer/dextramer-specific cells, a maxK of 10 and a random seed. Dimension reduction was performed using UMAP. Statistical comparison of marker expression levels was made using Kruskal-Wallis, with t tests for individual comparisons, and adjusted *P* values displayed. All code and FCS files used to undertake the analysis can be found at https://github.com/RebeccaPPayne/Omicron---Sheffield and https://github.com/RebeccaPPayne/Omicron---Newcastle.

## Supporting information

PITCH consortium author list

## Data Availability

All code and data used to undertake the Uniform Manifold Approximation and Projection (UMAP) analysis can be found on Github. An anonymised copy of all data used in this study will be made available on the Open Science Framework.

https://github.com/RebeccaPPayne/Omicron---Sheffield

https://github.com/RebeccaPPayne/Omicron---Newcastle

## Author contributions

Conceptualization: TdS, SJD, LT, PK, CJAD, RPP, SLRJ, MC, AR, EB

Methodology: TdS, SJD, LT, PK, CJAD, RPP, MC, AR, EB, SL, HH, GS

Formal analysis: HH, ARN, SL, CL, AT, AA, BK, MGR, TdS, RPP

Investigation: HH, ARN, SL, CL, AT, AA, BK, JKT, TT, PZ, MGR, PS, MS, PA, IN, MA, NAB, JMN, LG, SS, IG, TR, SCM, LMH, SLD, SB

Resources: AB, LT, EB

Data curation: HH, TdS, RPP

Writing – original draft: HH, TdS

Writing – Review and Editing: HH, ARN, SL, AA, NAB, SB, TD, EB, LT, SLRJ, MC, CJAD, PK, SJD, RPP, TdS

Visualization: HH, AA, ARN, RPP, TdS

Supervision: CL, BK, AA, TD, JM, TL, VH, SH, EB, GS, AR, LT, SLRJ, MC, CJAD, PK, SJD, RPP, TdS

Funding acquisition: PK, SJD, LT, TdS, CJAD, AR, SH, VH

## Acknolwedgements

This work was funded by the UK Department of Health and Social Care as part of the PITCH (Protective Immunity from T cells to Covid-19 in Health workers) Consortium, UKRI as part of “ Investigation of proven vaccine breakthrough by SARS-CoV-2 variants in established UK healthcare worker cohorts: SIREN consortium & PITCH Plus Pathway” MR/W02067X/1 and UKRI as part of “ PITCH2 - Protective Immunity through T Cells in Healthcare workers 2” MR/X009297/1, with contributions from UKRI/NIHR through the UK Coronavirus Immunology Consortium (UK-CIC), the Huo Family Foundation and The National Institute for Health Research (UKRIDHSC COVID-19 Rapid Response Rolling Call, Grant Reference Number COV19-RECPLAS).

EB. and PK are NIHR Senior Investigators and P.K. is funded by WT109965MA. SJD is funded by an NIHR Global Research Professorship (NIHR300791). RPP is funded by a Career Re-entry Fellowship (204721/Z/16/Z). CJAD was supported by fellowships from Wellcome (211153/Z/18/Z) and the Medical Research Council (MR/X001598/1). This study was supported by the NIHR Newcastle Clinical Research Facility. TD., JM. and GS are funded by the Chinese Academy of Medical Sciences (CAMS) Innovation Fund for Medical Science (CIFMS), China (grant number: 2018-I2M-2-002), Schmidt Futures, the Red Avenue Foundation and the Oak Foundation. The Wellcome Centre for Human Genetics is supported by the Wellcome Trust (grant 090532/Z/09/Z). LT is supported by the Wellcome Trust (grant number 205228/Z/16/Z) and the National Institute for Health Research Health Protection Research Unit (NIHR HPRU) in Emerging and Zoonotic Infections (NIHR200907) at University of Liverpool in partnership with UK Health Security Agency (UKHSA), in collaboration with Liverpool School of Tropical Medicine and the University of Oxford. DGW is supported by an NIHR Advanced Fellowship in Liverpool. MC, SL, LT, and TT are supported by U.S. Food and Drug Administration Medical Countermeasures Initiative contract 75F40120C00085. The Sheffield Teaching Hospitals Observational Study of Patients with Pulmonary Hypertension, Cardiovascular and other Respiratory Diseases (STH-ObS) was supported by the British Heart Foundation (PG/11/116/29288). The STH-ObS Chief Investigator Allan Laurie is supported by a British Heart Foundation Senior Basic Science Research fellowship (FS/18/52/33808). We gratefully acknowledge financial support from the UK Department of Health and Social Care via the Sheffield NIHR Clinical Research Facility award to the Sheffield Teaching Hospitals Foundation NHS Trust. The Wellcome Trust grant is acknowledged as UNS104697 Spectral Cytometry for profiling single cells of the immune system in health and disease to A Filby/NUFCCF.

The views expressed are those of the author(s) and not necessarily those of the NHS, the NIHR, the Department of Health or Public Health England.

## SUPPLEMENTARY INFORMATION

**Table S1.** Antibodies used for multiparameter flow cytometry (Newcastle)

| Reagent<br>(clone) | Source | Identifier | Titration | Reference<br>Control | Events<br>Acquired for<br>reference<br>control |
| --- | --- | --- | --- | --- | --- |
| CD62L-BB515<br>(SK11) | Becton<br>Dickinson | 565037 | 1/100 | Cells | 50,000 |
| CCR7-PerCP<br>(G043H7) | BioLegend | 353242 | 1/20 | Cells | 70,000 |
| CD57-PerCP_<br>Cy5.5 (HNK4) | BioLegend | 359622 | 1/100 | Cells | 60,000 |
| CD38-PerCP_<br>eFluor_710<br>(HB7) | ThermoFisher | 46-0388-42 | 1/50 | Cells | 50,000 |
| CD25-PE_Cy7<br>(M-A251) | Becton<br>Dickinson | 557741 | 1/50 | Cells | 50,000 |
| CD45RA-Alexa<br>Fluor 647<br>(HI100) | BioLegend | 304154 | 1/50 | Cells | 50,000 |
| CCR5-Alexa<br>Fluor 700<br>(HEK_1_85a) | BioLegend | 313713 | 1/100 | Cells | 100,000 |
| CD69-APC_Cy7<br>(FN50) | BioLegend | 310914 | 1/100 | Cells | 50,000 |
| CD3-APC Fire<br>810 (SK7) | BioLegend | 344858 | 1/200 | Cells | 30,000 |
| PD-1-BV421<br>(EH12.1) | Becton<br>Dickinson | 562516 | 1/50 | Cells | 50,000 |
| CD27-Super<br>Bright 436<br>(0323) | ThermoFisher | 62-0279-42 | 1/100 | Cells | 50,000 |
| CD45RO-Pacific<br>Blue (UCHL1) | BioLegend | 304216 | 1/100 | Cells | 50,000 |
| CCR2-BV480<br>(LS132.1D9) | Becton<br>Dickinson | 747852 | 1/100 | Cells | 100,000 |
| CD4-eFluor 506<br>(RPA-T4) | ThermoFisher | 69-0049-42 | 1/200 | Cells | 30,000 |
| HLA-DR-BV570<br>(L243) | BioLegend | 307638 | 1/50 | Cells | 50,000 |
| CD28-BV605<br>(L293) | Becton<br>Dickinson | 742527 | 1/50 | Cells | 50,000 |
| CD95-BV650<br>(DX2) | Becton<br>Dickinson | 740589 | 1/100 | Cells | 50,000 |
| KLRG1-BV711<br>(2F1) | BioLegend | 138427 | 1/50 | Cells | 80,000 |
| CXCR6-BV786<br>(13B 1E5) | Becton<br>Dickinson | 743602 | 1/20 | Cells | 150,000 |
| CD19-BUV395<br>(SJ25C1) | Becton<br>Dickinson | 563549 | 1/100 | Cells | 50,000 |
| CD127-BUV737<br>(HIL-7R-M21) | Becton<br>Dickinson | 612794 | 1/50 | Cells | 80,000 |
| CD8-BUV805<br>(SK1) | Becton<br>Dickinson | 359622 | 1/200 | Cells | 30,000 |
| **Surrogates** |  |  |  |  |  |
| CD3-APC<br>(UCHT1) | BioLegend | 300412 | 1/200 | Cells | 30,000 |
| CD3-PE<br>(UCHT1) | BioLegend | 300408 | 1/200 | Cells | 30,000 |

**Table S2.**
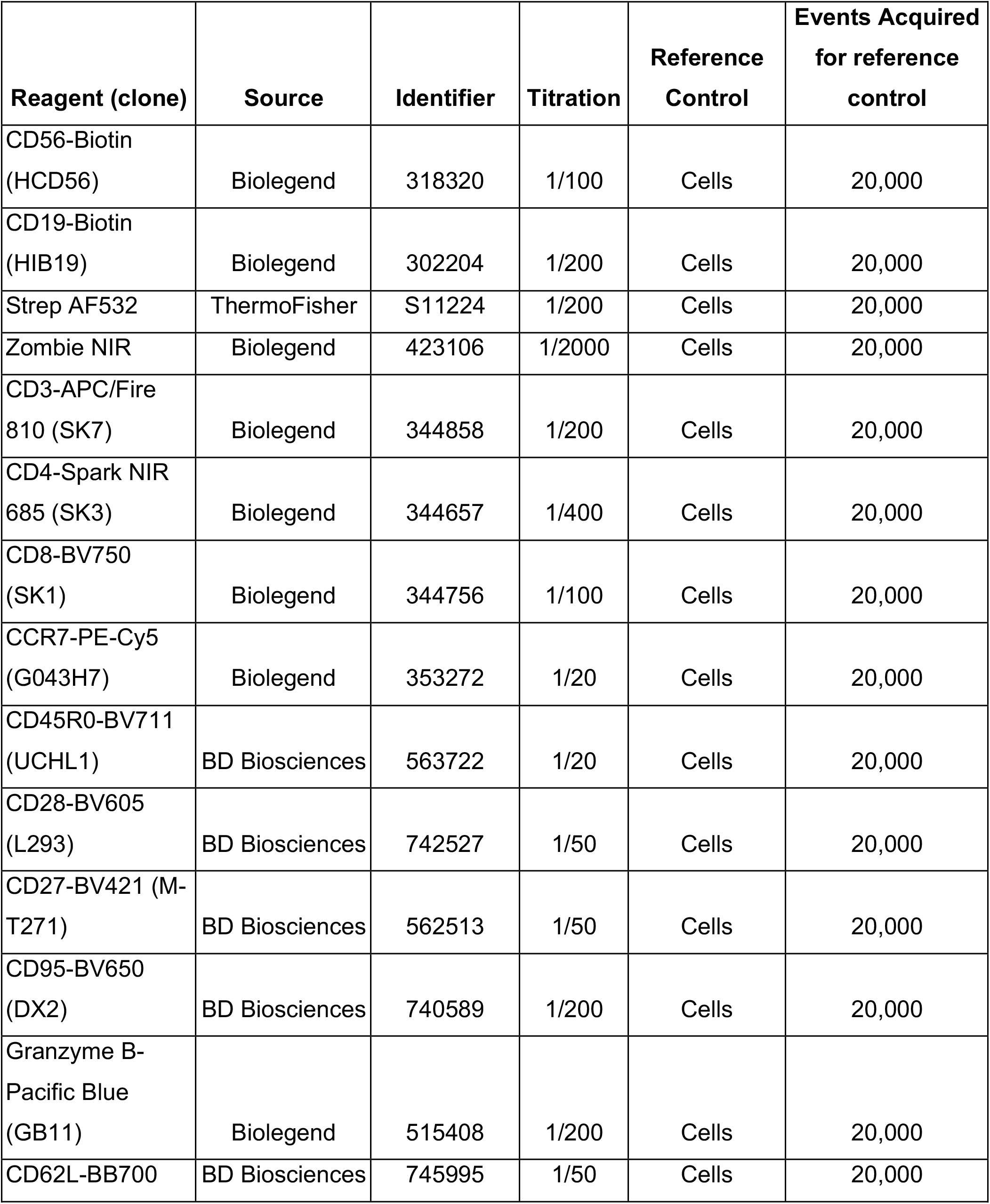
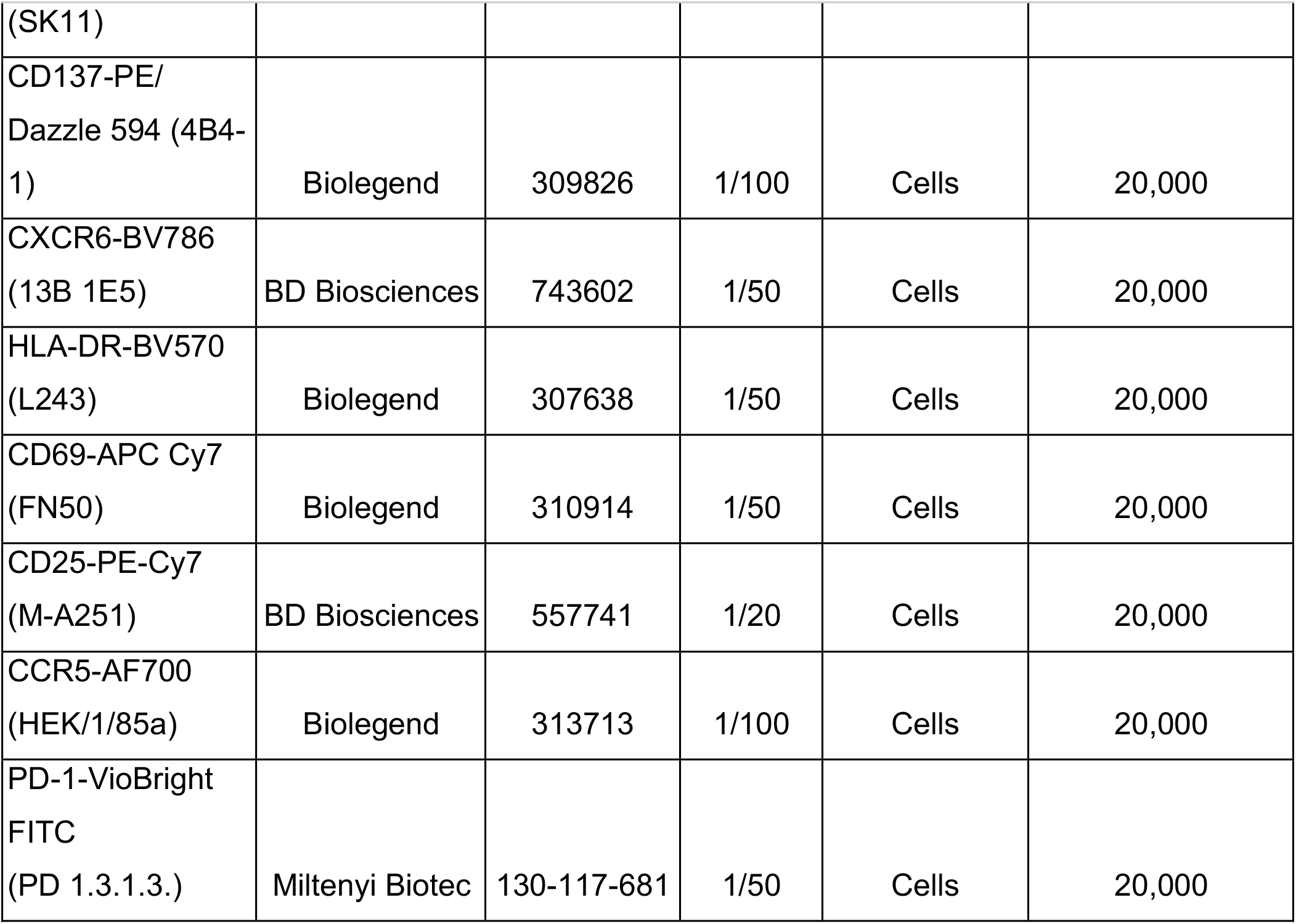
Antibodies used for multiparameter flow cytometry (Sheffield)

**Table S3.**
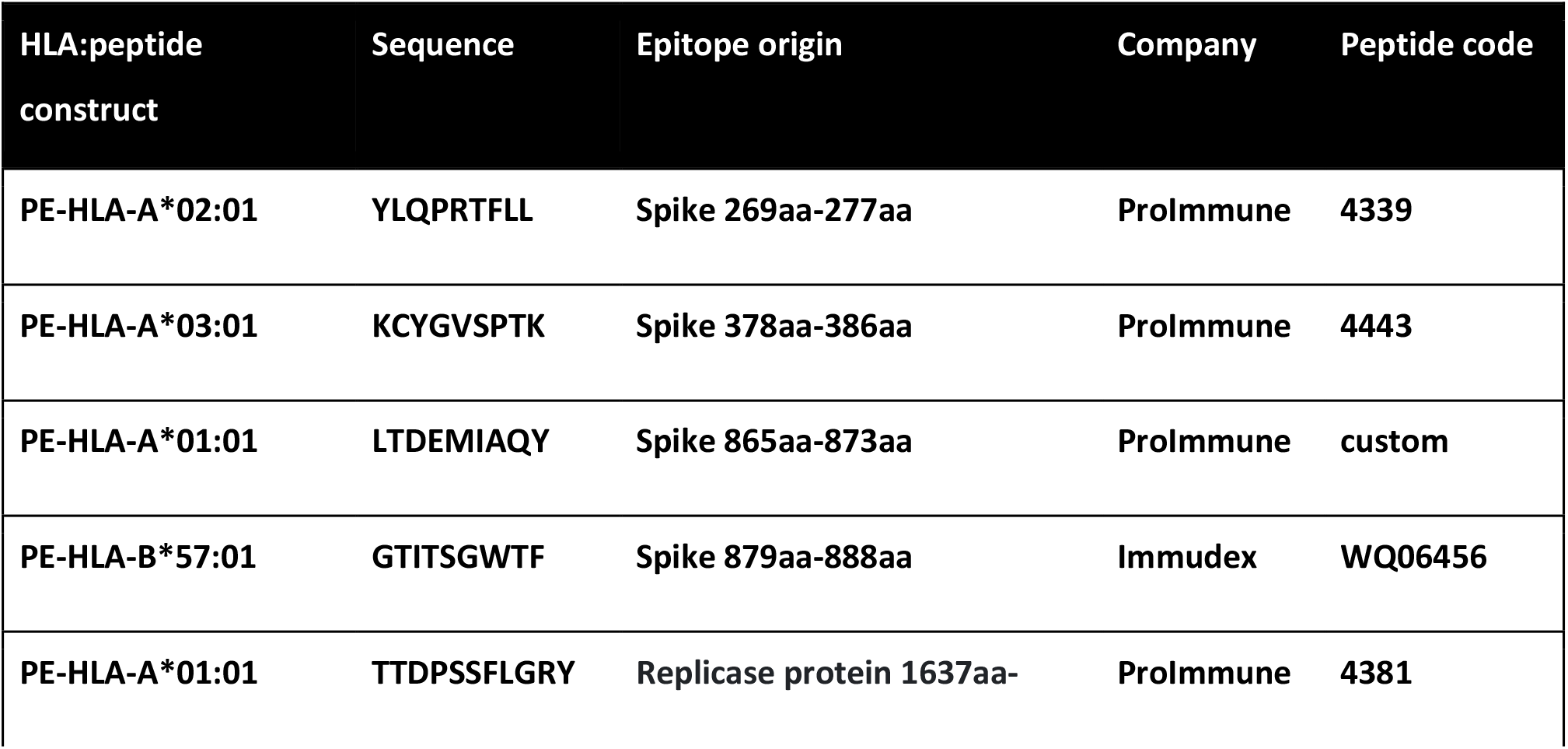
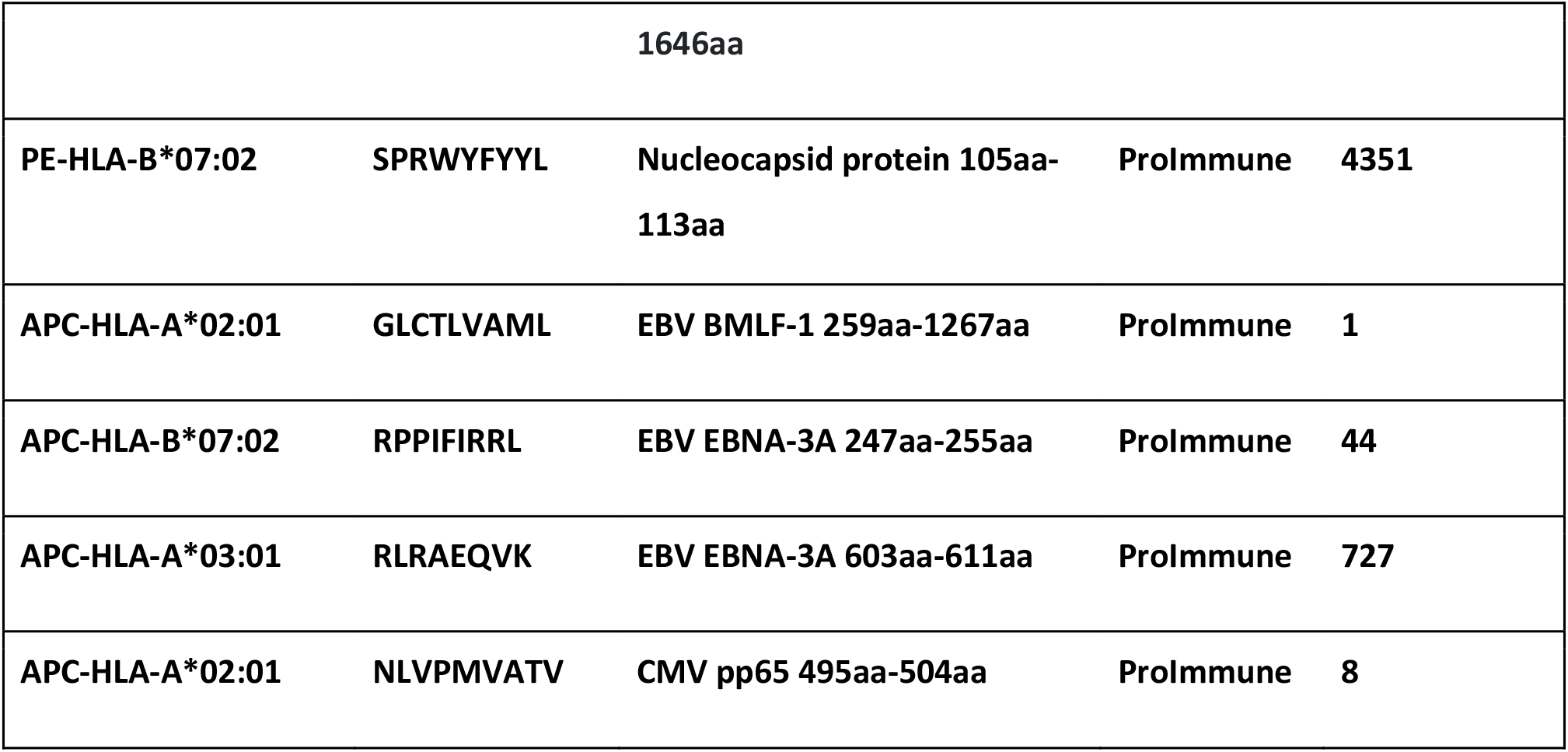
Pentamer/dextramer constructs used for staining the antigen-specific CD3+CD8+ T cells

**Figure S1.**
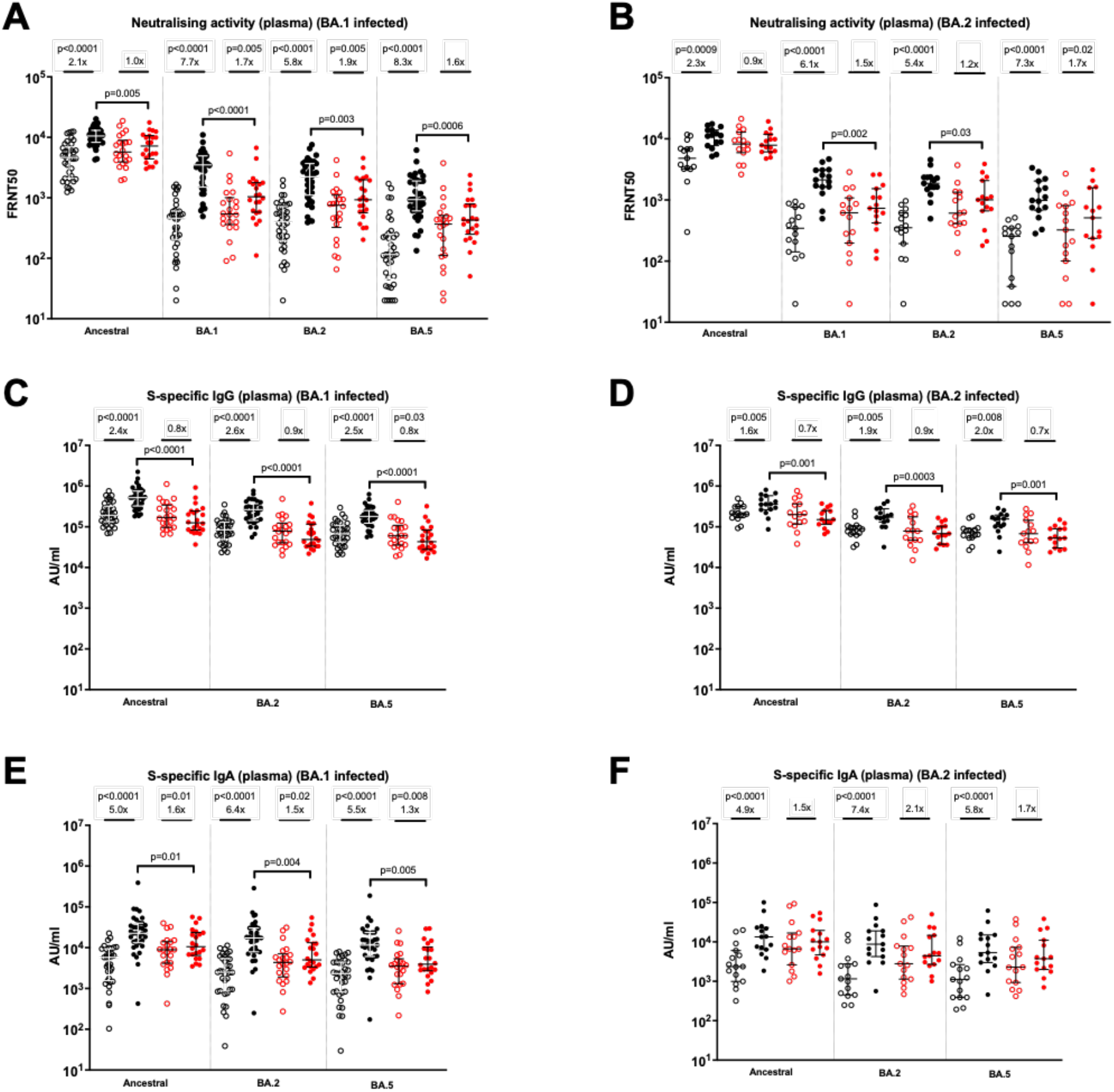
Impact of omicron infection on plasma neutralizing and binding antibodies in SARS-CoV-2-naive and previously-infected individuals, stratified by likely BA.1 or BA.2 infected individuals. Live-virus neutralizing activity of plasma against ancestral, BA.1, BA.2 and BA.5 viruses, expressed as the reciprocal of the dilution showing 50% reduction in focus forming units (FRNT50) in BA.1- (A) and BA.2- (B) infected individuals; SARS-CoV-2 spike-specific binding IgG in plasma against ancestral, BA.2 and BA.5 spike proteins (AU/mL = arbitrary antibody units/mL in MSD assay) in BA.1- (C) and BA.2- (D) infected individuals; SARS-CoV-2 spike-specific binding IgA in plasma against ancestral, BA.2 and BA.5 spike proteins (AU/mL = arbitrary antibody units/mL in MSD assay) in BA.1- (C) and BA.2- (D) infected individuals; Data shown with median and interquartile range. Median fold-change from pre-to post-infection samples is displayed. Statistical comparisons of pre- and post-infection samples done with Wilcoxon signed-rank test, and between post-infection levels in previously-infected and SARS-CoV-2 naive individuals using the Mann-Whitney U test. P values are displayed where <0.05. Responses were evaluated in 53 SARS-CoV-2-naive and 37 previously-infected individuals for whom samples were available.

**Figure S2.**
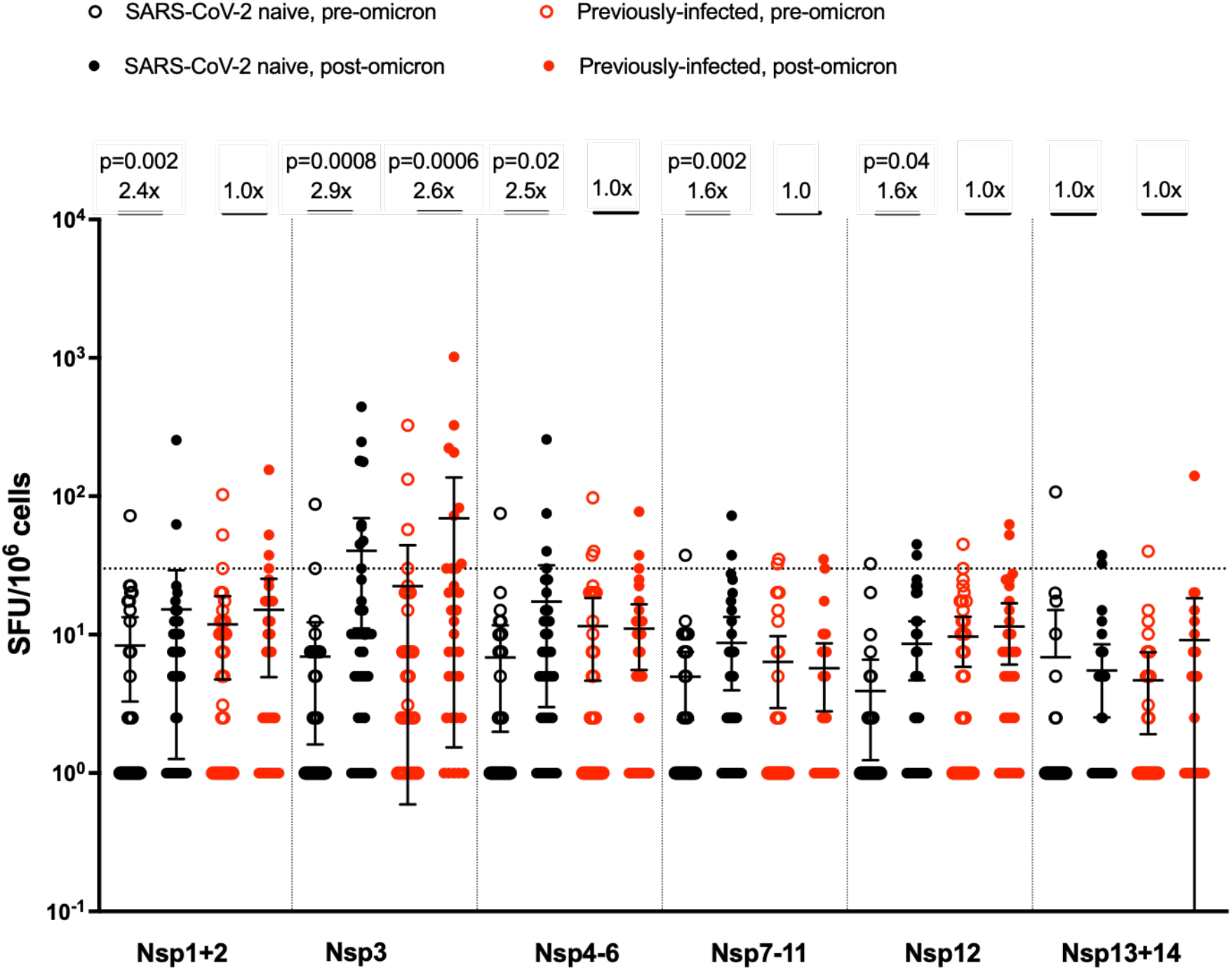
Impact of omicron infection on peptides representing non-structural proteins (NSP). IFN-**γ** ELISpot responses to overlapping peptide pools representing NSP 1-2, NSP3, NSP4-6, NSP7-11, NSP12 and NSP13 and 14. Results expressed as spot-forming units per million cells (SFU/106). The dashed line represents a positivity threshold of the mean + 2SD of the background response. Data shown with median and interquartile range. Median fold-change from pre-to post-infection samples is displayed. Statistical comparisons of pre- and post-infection samples done with Wilcoxon signed-rank test. P values are displayed where <0.05. Responses were evaluated in 37 SARS-CoV-2-naive and 32 previously-infected individuals for whom samples were available.

**Figure S3.**
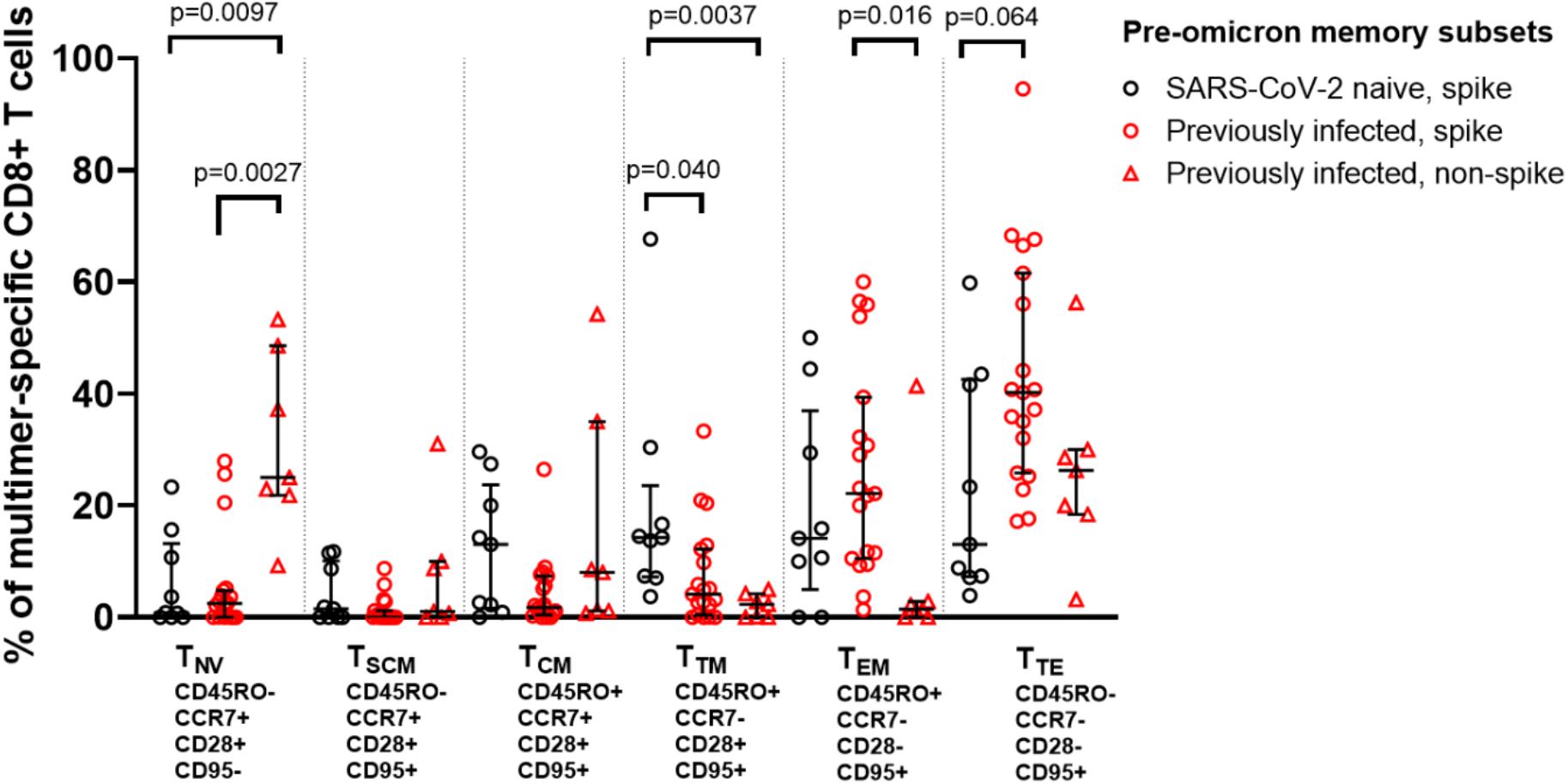
Epitope-specific CD8+ T cells phenotype following 3rd BNT162b2 mRNA vaccine dose but prior to omicron infection. Memory phenotypes of SARS-CoV-2-specific CD8+ T cells prior to omicron infection (pooled data from Sheffield and Newcastle). Shown are 9 spike-specific populations from 8 SARS-CoV-2 naive individuals, 19 spike-specific populations from 14 previously-infected individuals, and 8 non-spike populations from 7 previously-infected individuals. Data shown with median and interquartile range. Statistical comparisons between different multimer-specific populations were performed using Kruskal-Wallis test and Dunn’s post-hoc test for multiple pairwise comparisons. P values >0.05 unless displayed. T_NV_ = naive T cells, T_SCM_ = stem cell memory T cells, T_CM_ = central memory T cells, T_TM_ = transitional memory T cells, T_EM_ = effector memory T cells, T_TE_ = terminal effector T cells.

**Figure S4.**
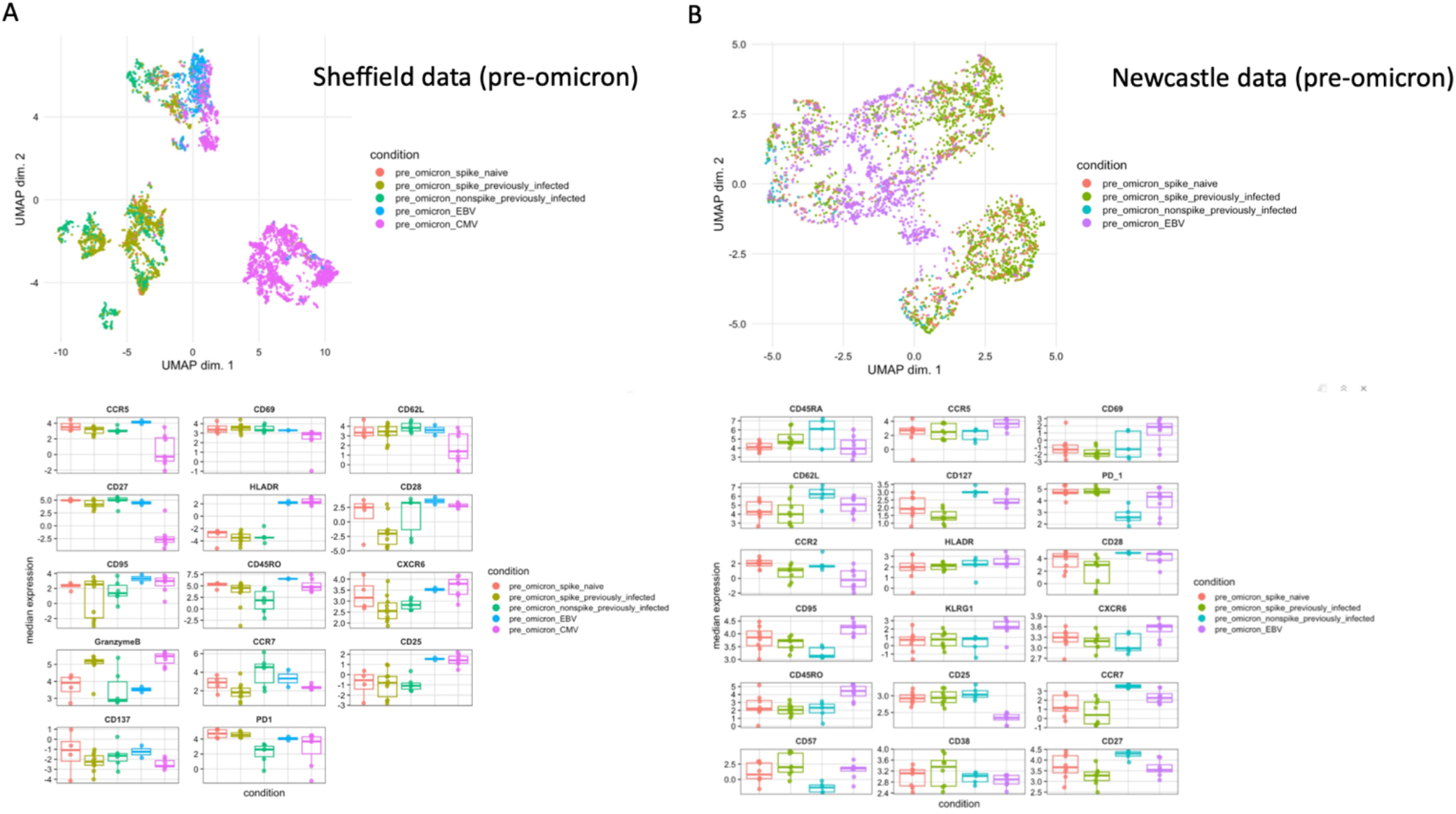
Phenotype of epitope-specific CD8+ T cell populations after 3^rd^ mRNA vaccine dose but prior to omicron infection, characterized using multi-dimensional flow cytometry. Uniform Manifold Approximation and Projection (UMAP) plots and marker expression levels showing multimer-specific cell clusters from SARS-CoV-2 spike-, SARS-CoV-2 non-spike-, EBV- and CMV-specific CD8+ T cells prior to omicron infection, using expression of 14 (Sheffield – A) or 18 (Newcastle - B) markers. CMV-specific populations were included in the Sheffield dataset only.

**Figure S5.**
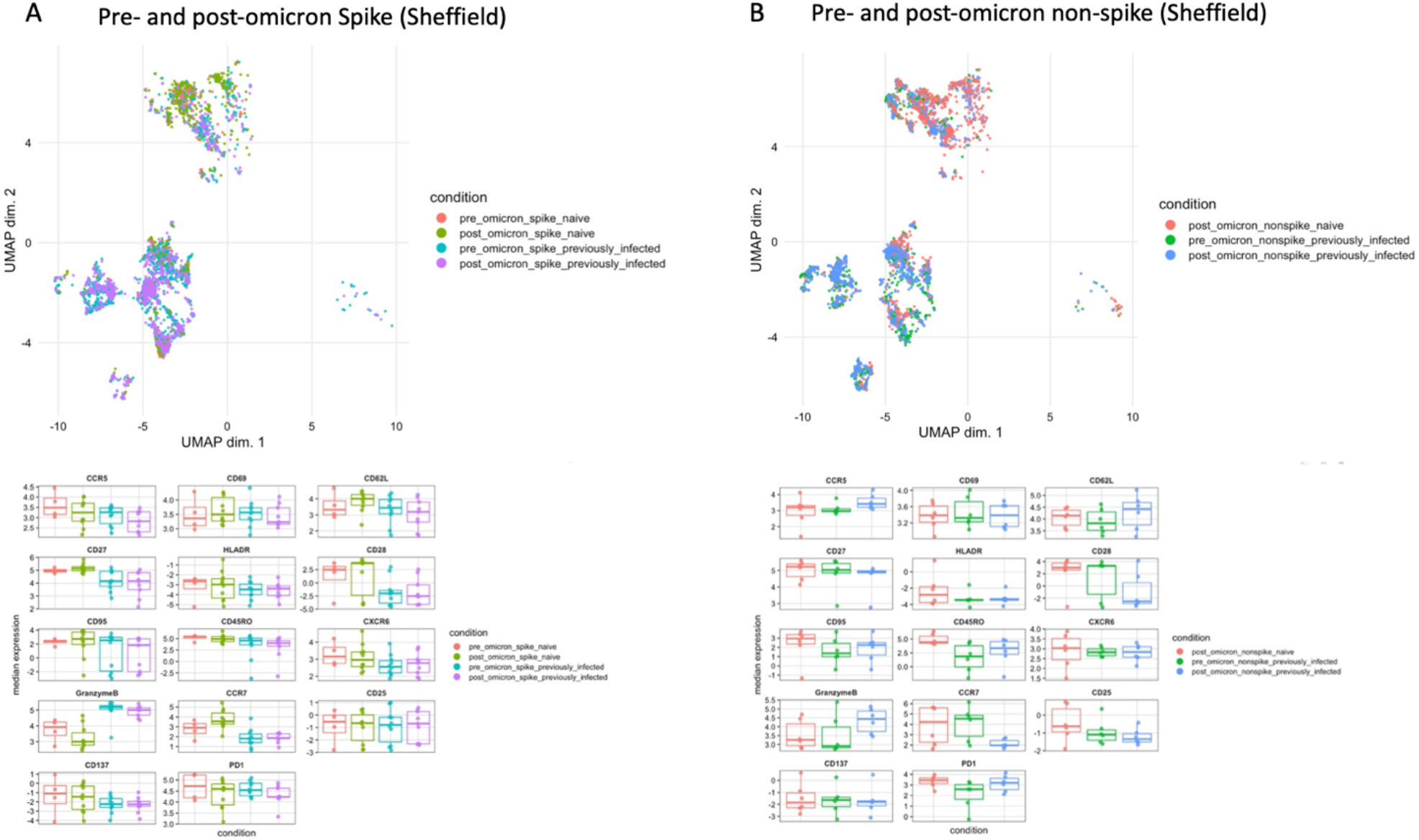
UMAP plots and expression of markers used in multi-dimensional flow cytometry in participants from Sheffield before and after omicron infection. Shown are data from (A) spike and (B) non-spike multimer-specific populations.

**Figure S6.**
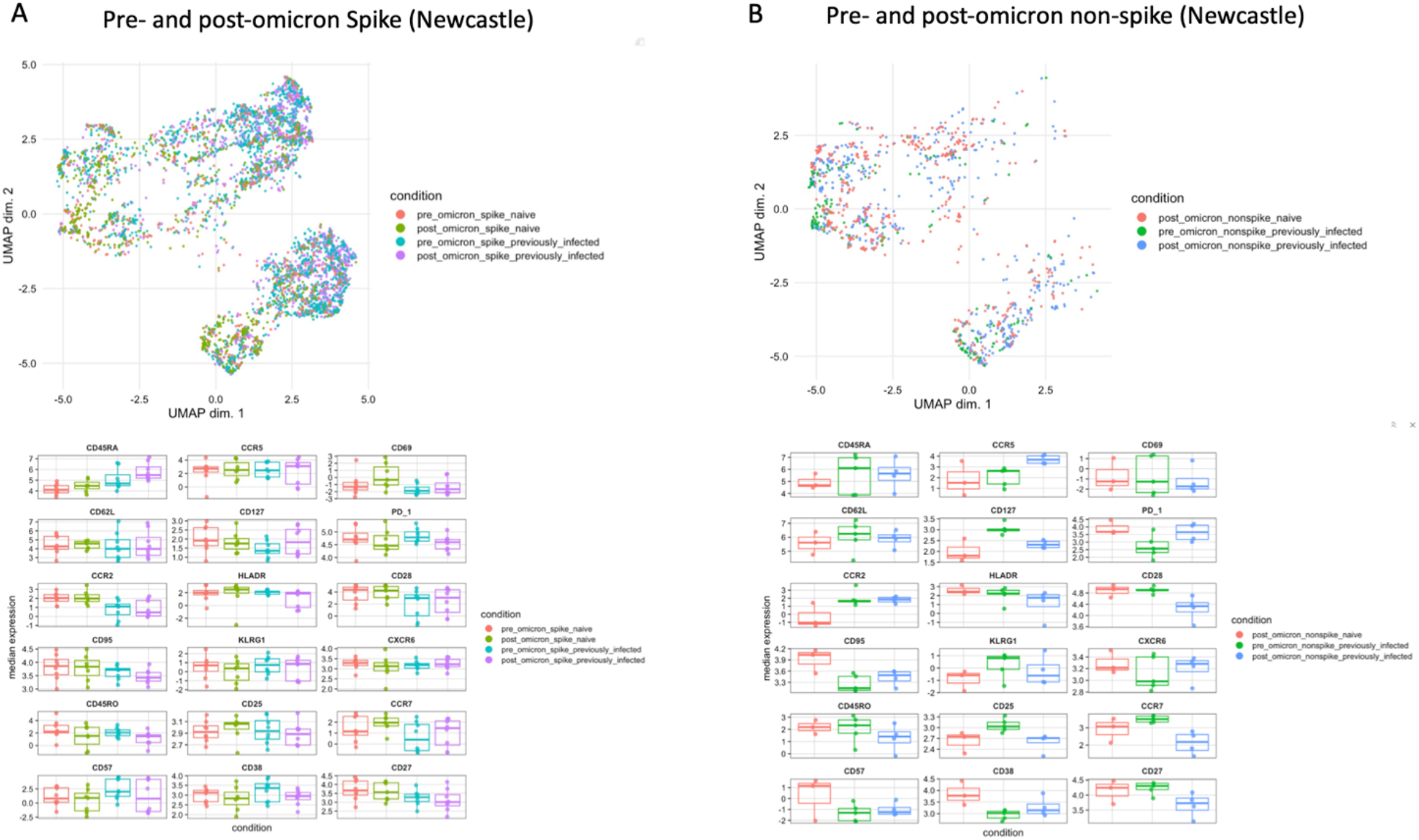
UMAP plots and expression of markers used in multi-dimensional flow cytometry in participants from Newcastle before and after omicron infection. Shown are data from (A) spike and (B) non-spike multimer-specific populations.

**Figure S7.**
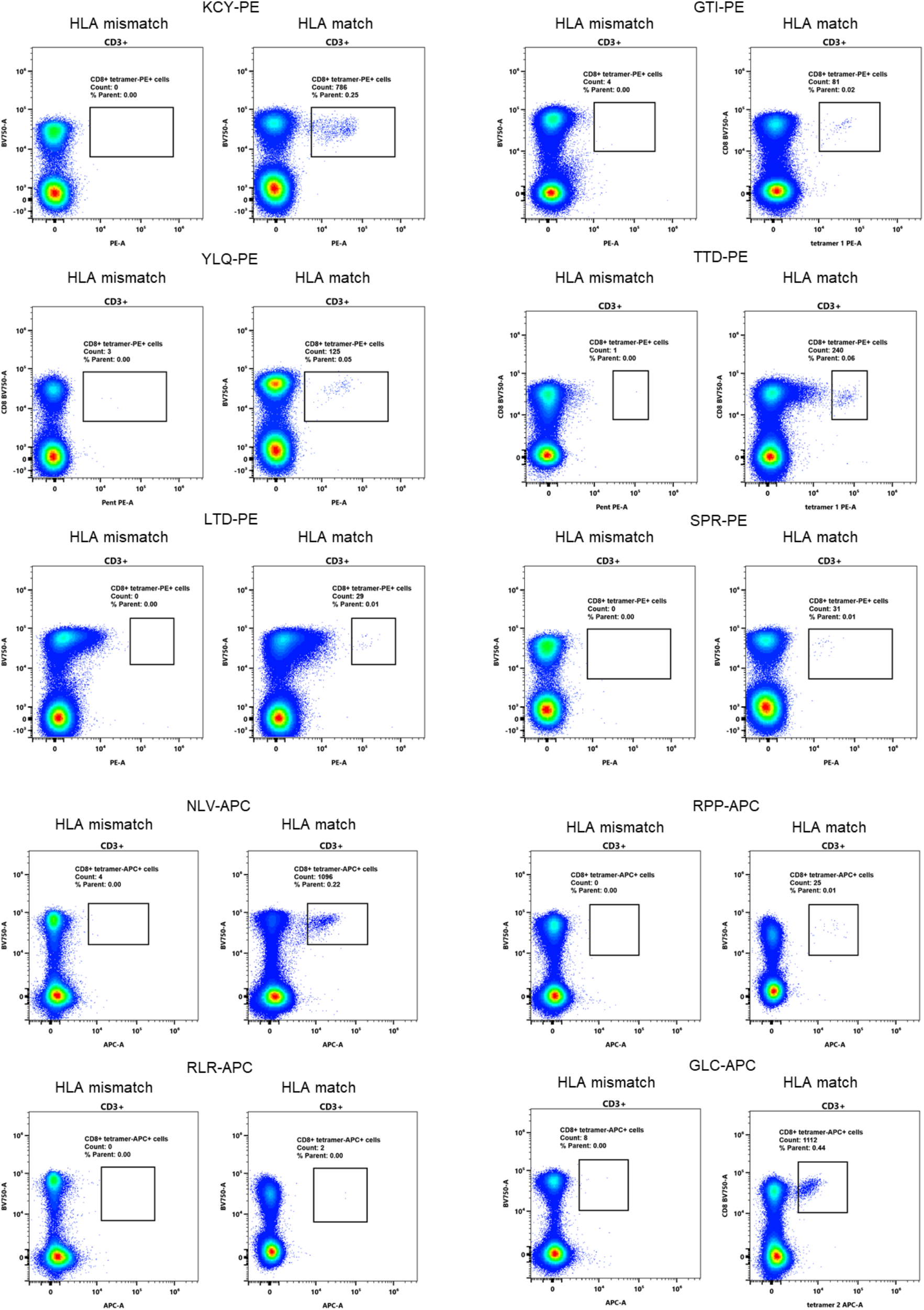
Examples of gating for multimer-specific CD8+ T cells. Shown are all multimer-specific populations included in the study (PE- or APC-labelled, X axis; CD8-on Y axis, after gating on CD3+ live, singlet cells), alongside staining of peripheral blood mononuclear cells from HLA-mismatched donors.

