## Supplementary material for "Omicron BA.1/BA.2 infections in triple-vaccinated individuals enhance a diverse repertoire of mucosal and blood immune responses": PITCH consortium author list

| First Name | | Last Name | Institution |
|---|---|---|---|
| Jenna | | Ablott | Sheffield Teaching Hospitals NHS Foundation Trust |
| Priyanka | | Abraham | University of Oxford |
| Sandra | | Adele | University of Oxford |
| Zahra | | Ahmed | University of Birmingham |
| Saly | | Al-Taei | University of Birmingham |
| Mohammad | | Ali | University of Oxford |
| Ali | | Amini | University of Oxford |
| Adrienn | | Angyal | University of Sheffield |
| M. | Azim | Ansari | University of Oxford |
| Rachel | | Anslow | University of Oxford |
| Ana | | Atti | UK Health Security Agency |
| James | | Austin | University of Liverpool |
| Angela | | Bailey | Newcastle University |
| Eleanor | | Barnes | University of Oxford |
| Natalie | A. | Barratt | University of Sheffield |
| Martin | | Bayley | University of Sheffield |
| Sagida | | Bibi | University of Oxford |
| Lucy | H. | Booth | University of Cambridge |
| Alice | | Bridges-Webb | University of Oxford |
| Anthony | | Brown | University of Oxford |
| Rebecca | | Brown | University of Sheffield |
| Holly | | Caborn | Sheffield Teaching Hospitals NHS Foundation Trust |
| Miles | | Carroll | University of Oxford |
| Jeremy | | Chalk | University of Oxford |
| Anu | | Chawla | Liverpool University Hospitals NHS Foundation Trus |
| Elizabeth | | Clutterbuck | University of Oxford |
| Christopher | P. | Conlon | University of Oxford |
| Andrew | | Cross | Liverpool University Hospitals NHS Foundation Trus |
| Debbie | | Cross | University of Oxford |
| Sophie | | Davies | University of Oxford |
| Catherine | | de Lara | University of Oxford |
| Thushan | I. | de Silva | University of Sheffield |
| Alexandra | S | Deeks | University of Oxford |
| Wanwisa | | Dejnirattisai | University of Oxford |
| Susan | L | Dobson | University of Liverpool |
| Christina | | Dold | University of Oxford |
| Thomas | M. | Drake | University of Edinburgh |
| Susanna | | Dunachie | University of Oxford |
| Christopher | JA | Duncan | Newcastle University |
| Elena | | Efstathiou | University of Birmingham |
| David | | Eyre | University of Oxford |
| Alex | | Fairman | University of Sheffield |
| Sian | | Faustini | University of Birmingham |
| Andrew | | Filby | Newcastle University |
| Sarah | | Foulkes | UK Health Security Agency |
| John | | Frater | University of Oxford |
| Lisa | | Frending | University of Oxford |
| Oliver | | Galgut | University of Birmingham |
| Siobhan | | Gardiner | University of Oxford |
| Philip | | Goulder | University of Oxford |
| Jessica | | Gregory | Sheffield Teaching Hospitals NHS Foundation Trust |

| | | | |
|---|---|---|---|
| Irina | | Grouneva | University of Sheffield |
| Lotta | | Gustafsson | Sheffield Teaching Hospitals NHS Foundation Trust |
| Carl-Philipp | | Hackstein | University of Oxford |
| Victoria | | Hall | UK Health Security Agency |
| Callum | | Halstead | University of Oxford |
| Sophie | | Hambleton | Newcastle University |
| Muzlifah | | Haniffa | Newcastle University |
| Helen | | Hanson | Newcastle University |
| Alexander | | Hargreaves | University of Oxford |
| Kate | | Harrington | Sheffield Teaching Hospitals NHS Foundation Trust |
| Jenny | | Haworth | Newcastle upon Tyne Hospitals NHS Foundation Trust |
| Carole | | Hays | Newcastle University |
| Luisa | M | Hering | University of Liverpool |
| Susan | | Hopkins | UK Health Security Agency |
| Emily | C. | Horner | University of Cambridge |
| Hailey | | Hornsby | University of Sheffield |
| Fatima | Mari | Ilyas | Sheffield Teaching Hospitals NHS Foundation Trust |
| Jasmin | | Islam | UK Health Security Agency |
| Anni | | Jämsén | University of Oxford |
| Katie | | Jeffery | University of Oxford |
| Sile | | Johnson | University of Oxford |
| Geraldine | | Jones | Newcastle University |
| Mwila | | Kasanyinga | University of Oxford |
| Sinead | | Kelly | Newcastle upon Tyne Hospitals NHS Foundation Trust |
| Maqsood | | Khan | Sheffield Teaching Hospitals NHS Foundation Trust |
| Jon | | Kilby | University of Sheffield |
| Rosemary | | Kirk | Sheffield Teaching Hospitals NHS Foundation Trust |
| Paul | | Klenerman | University of Oxford |
| Barbara | | Kronsteiner | University of Oxford |
| Teresa | | Lambe | University of Oxford |
| Allan | | Lawrie | University of Sheffield |
| Lauren | | Lett | University of Liverpool |
| Chang | | Liu | University of Oxford |
| Stephanie | | Longet | University of Oxford |
| Alison | | Lye | Sheffield Teaching Hospitals NHS Foundation Trust |
| Tom | | Malone | University of Oxford |
| Spyridoula | | Marinou | University of Oxford |
| Chloe | | Matthewman | Sheffield Teaching Hospitals NHS Foundation Trust |
| Philippa | C. | Matthews | Francis Crick Institute |
| David | | McDonald | Newcastle University |
| Jessica | | McNeill | Sheffield Teaching Hospitals NHS Foundation Trust |
| Gracie | | Mead | University of Oxford |
| Naomi | | Meardon | Sheffield Teaching Hospitals NHS Foundation Trust |
| Alexander | J. | Mentzer | University of Oxford |
| Shagun | | Misra | Sheffield Teaching Hospitals NHS Foundation Trust |
| Juthathip | | Mongkolsapaya | University of Oxford |
| Shona | C | Moore | University of Liverpool |
| Sam | M. | Murray | University of Oxford |
| Isabel | | Neale | University of Oxford |
| Jeremy | M. | Nell | Newcastle University |
| Thomas | AH | Newman | Sheffield Teaching Hospitals NHS Foundation Trust |
| Alexander | R | Nicols | Newcastle University |

| | | | |
|---|---|---|---|
| Christopher | | Norman | Sheffield Teaching Hospitals NHS Foundation Trust |
| Ane | | Ogbe | University of Oxford |
| Ashley | | Otter | UK Health Security Agency |
| Juyeon | | Park | University of Oxford |
| Brendan | A.I. | Payne | Newcastle University |
| Rebecca | P. | Payne | Newcastle University |
| Eloise | | Phillips | University of Oxford |
| Gareth | | Platt | University of Liverpool |
| Megan | | Plowright | Sheffield Teaching Hospitals NHS Foundation Trust |
| Andrew | J. | Pollard | University of Oxford |
| Sonia | | Poolan | Newcastle upon Tyne Hospitals NHS Foundation Trust |
| Nicholas | | Provine | University of Oxford |
| Alex Richter | | Richter | University of Birmingham |
| Chloe | | Roddis | Sheffield Teaching Hospitals NHS Foundation Trust |
| Stefan | | Roman | Sheffield Teaching Hospitals NHS Foundation Trust |
| Leigh | | Romaniuk | Newcastle upon Tyne Hospitals NHS Foundation Trust |
| Patpong | | Rongkard | University of Oxford |
| Sarah | L. | Rowland-Jones | University of Sheffield |
| Ayoub | | Saei | UK Health Security Agency |
| Jose | | Schutter | University of Sheffield |
| Gavin | | Screaton | University of Oxford |
| Adrian | | Shields | University of Birmingham |
| Laura | | Silva Reyes | University of Oxford |
| Donal | | Skelly | University of Oxford |
| Nikki | | Smith | University of Sheffield |
| Jarmila | S. | Spegarova | Newcastle University |
| Lizzie | | Stafford | University of Oxford |
| Gareth | | Stephens | Sheffield Teaching Hospitals NHS Foundation Trust |
| Emily | | Stephenson | Newcastle University |
| Rachel | | Stimpson | Sheffield Teaching Hospitals NHS Foundation Trust |
| Scarlett | | Strickland | Sheffield Teaching Hospitals NHS Foundation Trust |
| Krishanthi | | Subramaniam | University of Liverpool |
| Piyada | | Supasa | University of Oxford |
| Chloe | | Tanner | University of Birmingham |
| Lydia | J. | Taylor | Newcastle University |
| Chitra | | Tejpal | University of Oxford |
| James | E.D. | Thaventhiran | University of Cambridge |
| Nicola | | Tinker | Sheffield Teaching Hospitals NHS Foundation Trust |
| Tom | | Tipton | University of Oxford |
| Neal | | Townsend | University of Birmingham |
| Simon | | Travis | University of Oxford |
| Nicola | | Trewick | Newcastle University |
| Stephanie | | Tucker | Newcastle University |
| Aekkachai | | Tuekprakhon | University of Oxford |
| Lance | | Turtle | University of Liverpool |
| Helena | | Turton | University of Sheffield |
| Jessica | K | Tyerman | Newcastle University |
| Zara | | Valiji | University of Oxford |
| Lisa | | Watson | Sheffield Teaching Hospitals NHS Foundation Trust |
| Rachel | | Whitham | Sheffield Teaching Hospitals NHS Foundation Trust |
| Jayne | | Willson | Sheffield Teaching Hospitals NHS Foundation Trust |
| Barbara | | Wilson | Newcastle University |

| | | | |
|---|---|---|---|
| Joseph | D. | Wilson | University of Oxford |
| Steven | | Wood | University of Sheffield |
| Daniel | G. | Wootton | University of Liverpool |
| Amira | A. T. | Zawia | Sheffield Teaching Hospitals NHS Foundation Trust |
| Martha | | Zewdie | University of Oxford |
| Peijun | | Zhang | University of Sheffield |
